# A systematic review of interleukin-2-based immunotherapies in clinical trials for cancer and autoimmune diseases

**DOI:** 10.1101/2022.12.02.22283042

**Authors:** Miro E. Raeber, Dilara Sahin, Ufuk Karakus, Onur Boyman

**Affiliations:** Department of Immunology, University Hospital Zurich, Zurich, Switzerland; Faculty of Medicine and Faculty of Science, University of Zurich, Zurich, Switzerland

**Keywords:** interleukin 2, IL-2, cancer, autoimmune disease, immunotherapy.

## Abstract

**Background:** The cytokine interleukin-2 (IL-2) can stimulate both effector immune cells and regulatory T (Treg) cells. The ability of selectively engaging either of these effects has spurred interest in using IL-2 for immunotherapy of cancer and autoimmune diseases. Thus, numerous IL-2-based biologic agents with improved bias or delivery toward effector immune cells or Treg cells have been developed. These improved IL-2-based compounds recently entered clinical trials.

**Objective:** This study systematically reviews clinical results of improved IL-2-based compounds for the treatment of cancer or autoimmune diseases.

**Methods:** We followed the Preferred Reporting Items for Systematic Reviews and Meta-Analyses (PRISMA), searched the ClinicalTrials.gov database for registered IL-2 trials using improved IL-2-based agents and different databases for available results of these studies.

**Results:** We identified 547 registered clinical trials, of which we extracted 36 studies on improved IL-2-based compounds. Moreover, we assessed another 9 agents reported in two recent literature reviews and based on our knowledge, totaling in 45 improved IL-2-based compounds. A secondary search for registered clinical trials of each of these improved 45 compounds resulted in 139 clinical trials included in this systematic review, with 29 trials reporting clinical results.

**Conclusions:** As of yet, none of the improved IL-2-based compounds gained regulatory approval for the treatment of cancer or autoimmune diseases. Three compounds treating cancer have entered phase 3 trials with two studies still ongoing. NKTR-214 is the only compound that has completed phase 3 studies. The PIVOT IO-001 study testing the combination of NKTR-214 plus Pembrolizumab compared to Pembrolizumab monotherapy in metastatic melanoma missed its primary endpoint of superior objective response rate and progression-free survival. The PIVOT-09 study, combining NKTR-214 with Nivolumab compared to Sunitinib or Cabozantinib in advanced renal cell carcinoma, missed its primary endpoint of improved objective response rate. Trials in autoimmune diseases are currently in early stages, thus not allowing conclusions on efficacy. Results of ongoing trials will provide insight into which improved IL-2-based compounds will be beneficial for cancer and autoimmune diseases.

## 1. Introduction

IL-2 is a small 15-kDa cytokine with pleiotropic effects on the immune system. Low doses of IL-2 preferentially bind to the trimeric IL-2 receptor (IL-2R), consisting of IL-2Rα (CD25), IL-2Rβ (CD122), and the common gamma chain (CD132), which is mainly expressed on immunosuppressive regulatory T (Treg) cells. Trimeric IL-2Rs are also referred to as high-affinity IL-2Rs because their affinity for IL-2 is about 10– 100 times higher than that of dimeric IL-2Rs (*1*). Once the limited amounts of trimeric IL-2Rs on these cells are saturated, IL-2 also very efficiently associates with and stimulates dimeric IL-2R, made of CD122 and CD132, that are principally present on antigen-experienced (memory) resting effector T (Teff) and natural killer (NK) cells (**Fig. 1**) (1, 2). The stimulatory effect of IL-2 on Teff and NK cells spurred the development of high-dose IL-2 for the treatment of cancer, with recombinant human IL-2 (Aldesleukin) becoming the first US Food and Drug Administration approved immunotherapy for the treatment of metastatic renal cell carcinoma (RCC) and metastatic melanoma in 1992 and 1998, respectively (*3, 4*). However, the stimulatory effects of high-dose IL-2 on Treg cells that dampen immune responses against self-antigens including certain tumor antigens, as well as the considerable adverse side effects of IL-2 at high doses due to vascular leak syndrome limited its efficacy in cancer. Thus, only 9.3 % of patients with RCC and 4.0 % with metastatic melanoma achieved complete responses (CR) on high-dose IL-2 monotherapy (*5, 6*). These shortcomings of high-dose IL-2 treatment motivated the development of improved IL-2-based biologic agents with higher selectivity for certain immune cell subsets and reduced toxicity (*7*). Furthermore, due to the success of immune checkpoint inhibitors, including monoclonal antibodies (mAbs) directed against programmed cell death protein 1 (PD-1) and cytotoxic T-lymphocyte-associated protein 4 (CTLA-4), IL-2 immunotherapy regained the interest of the pharmaceutical industry. On this point, it is worth noting the complementary mode of action of checkpoint inhibitors and IL-2 immunotherapy. Whereas checkpoint inhibitors show limited efficacy in non-immunogenic and poorly immune cell-infiltrated tumors, IL-2 might render these tumors immunogenic and thus amenable to checkpoint inhibitor treatment due to indirect stimulation of and tumor infiltration by dendritic cells (*8*).

**Fig. 1.**
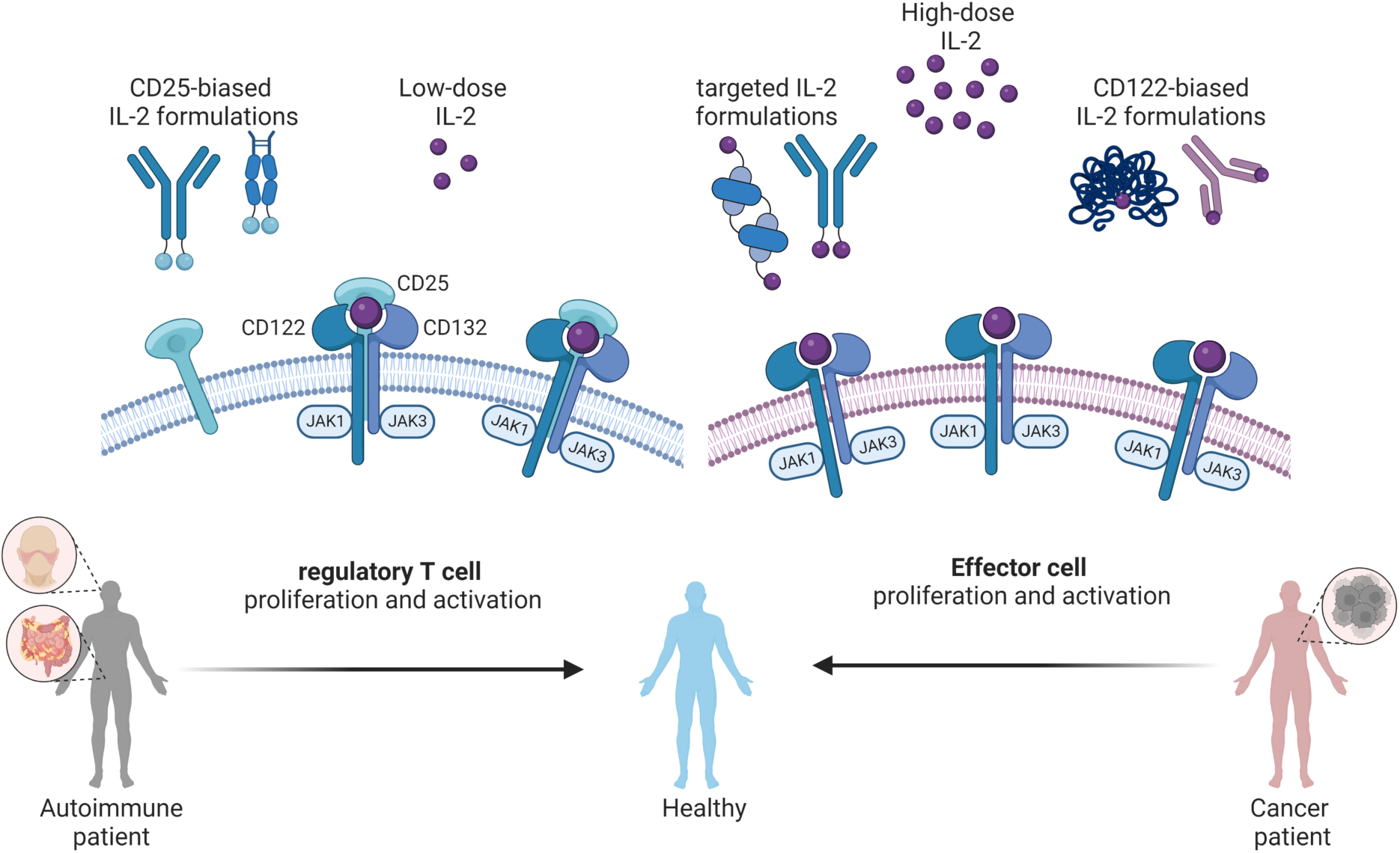
Biology of interleukin-2 (IL-2). Low-dose IL-2 and improved IL-2 formulations with CD25-bias preferentially stimulate the trimeric IL-2 receptor consisting of CD122, CD132, and CD25, thus expanding regulatory T (Treg) cells. Treg expansion restores immune balance in patients with autoimmune diseases such as systemic lupus erythematosus and inflammatory bowel diseases (left panel). On the other hand, high-doses of IL-2 or CD122-biased IL-2 formulations preferentially stimulate the dimeric IL-2 receptor consisting for CD122 and CD132 expressed on effector T and natural killer cells. Stimulation of these effector cells improves anti-tumor responses in cancer patients (right panel). Another approach focuses on delivery of IL-2 to either the tumor microenvironment or immune cells by using targeted IL-2 formulations (right panel).

The discovery of highly IL-2-dependent immunosuppressive Treg cells in 1995 and following research reporting Treg cell deficiencies in various autoimmune diseases prompted first clinical trials testing low-dose IL-2 immunotherapy for the treatment of chronic graft-versus-host disease and cryoglobulinemic vasculitis (*9–14*). Following these seminal trials, controlled clinical trials testing low-dose IL-2 in systemic lupus erythematosus (SLE) confirmed expansion of Treg cells by IL-2 immunotherapy, but these trials missed their predefined primary endpoints on clinical efficacy compared to control groups (*15, 16*). One explanation for this failure could be the imperfect bias of low-dose IL-2, which can stimulate also Teff and NK cells. This rationale has supported current endeavors of developing improved IL-2-based compounds with increased CD25 bias for selective activation and proliferation of Treg cells.

With registered clinical trials on improved IL-2-based agents increasing markedly during the past years (**Fig. 2**), the present study aims to provide a comprehensive review systemically summarizing our current knowledge on the available clinical efficacy data, compared to their molecular structure, of improved IL-2- based compounds that are currently in clinical trials.

**Fig. 2.**
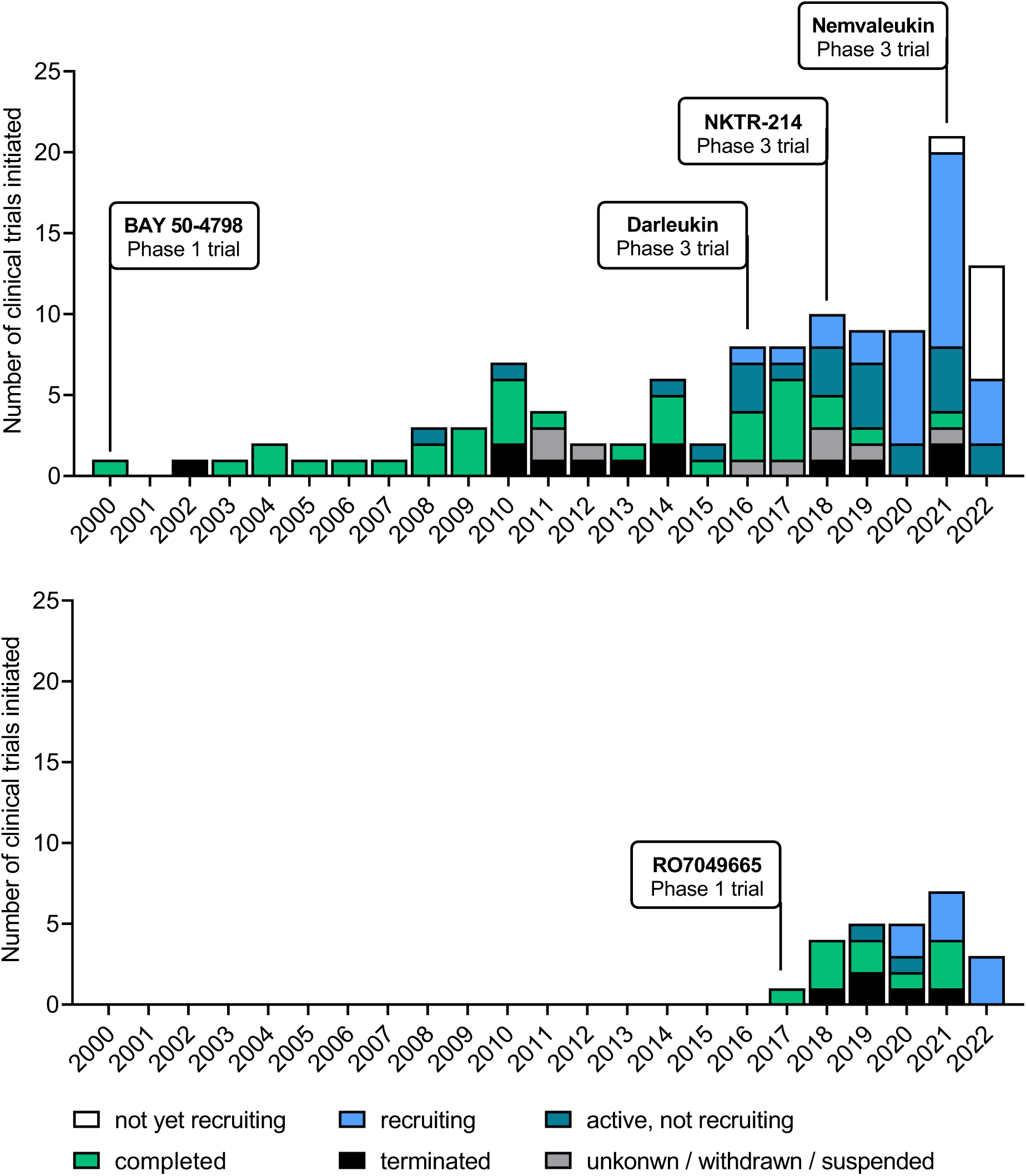
Registered clinical trials testing improved IL-2-based compounds. Top panel displays trials in cancer and bottom panel trials in autoimmune diseases.

**Fig. 3.**
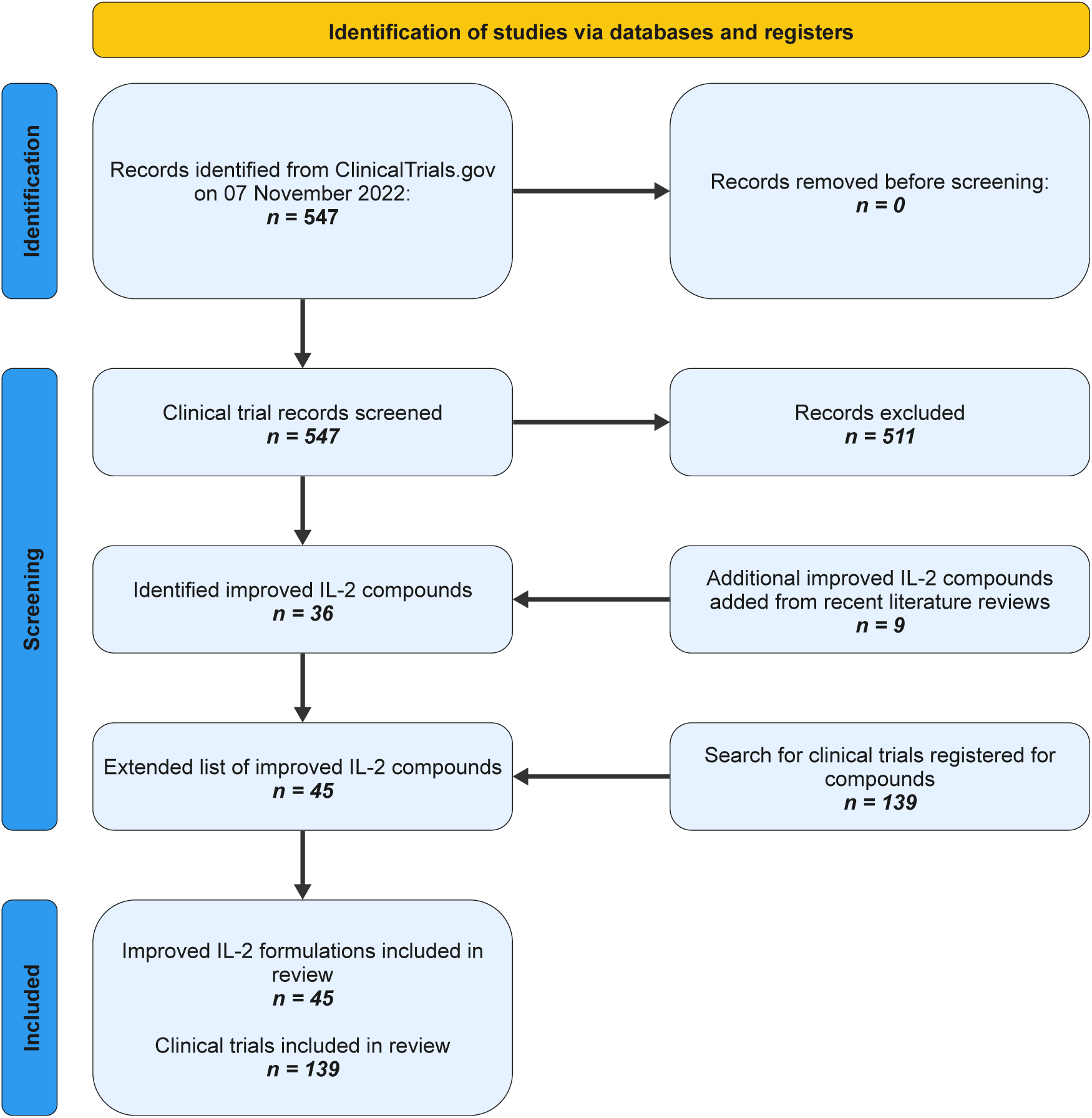
PRISMA flow-chart of literature research.

## 2. Methods

### Study design and protocol registration

This systematic review followed the Preferred Reporting Items for Systematic Reviews and Meta-Analyses (PRISMA) guidelines (**Table S1**) (*17*) and was registered with the International Platform of Registered Systematic Review and Meta-analysis Protocols (INPLASY) prior to the initiation of the literature search (Registration number: INPLASY2022110086).

### Search strategy

The ClinicalTrials.gov database was searched for registered clinical trials by applying in the field “Intervention/treatment” the search terms “interleukin-2” OR “interleukin 2” OR “IL-2” OR “IL2”. The results were filtered for clinical trials with study start between 01.01.2010 and 31.10.2022. The list of compounds was amended with missing IL-2-based formulations identified by recent literature reviews and based on the authors knowledge (*4, 18*).

### Eligibility criteria

Improved IL-2-based compounds in clinical development with registered clinical trials on ClinicalTrials.gov or reported results. Improved IL-2-based compounds were defined as IL-2-based biologic agents with a skewed bias toward or delivery to effector immune cells versus Treg cells compared to native IL-2. Clinical trials testing native IL-2, including Aldesleukin, or non-agonistic IL-2 formulations were excluded.

### Study selection, data collection process and analysis

Two authors (MR and UK) independently conducted the primary search and screened clinical trials testing improved IL-2-based compounds for inclusion. The resulting separate lists of clinical trials were compared, and improved IL-2-based compounds selected. In case of disagreement, a third author (DS) was involved in the discussion to reach final consensus of whether to include or not a given trial. To find all registered clinical trials for identified improved IL-2 compounds, a second round of search was performed for each of the identified compounds by using the ClinicalTrials.gov database, including alternative names of compounds. All identified clinical trials were subsequently listed in **Tables 1 and 2** and amended with available information on the clinical development status. For all studies, the trial phase, treatment arms and – where available – the results were retrieved from the ClinicalTrials.gov database. For completed trials with no results available on ClinicalTrials.gov, a targeted Medical Literature Analysis and Retrieval System Online (MEDLINE) and internet search was conducted to retrieve study results. Non-retrievable trial results were confirmed by an independent search by a second author.

**Table 1:**
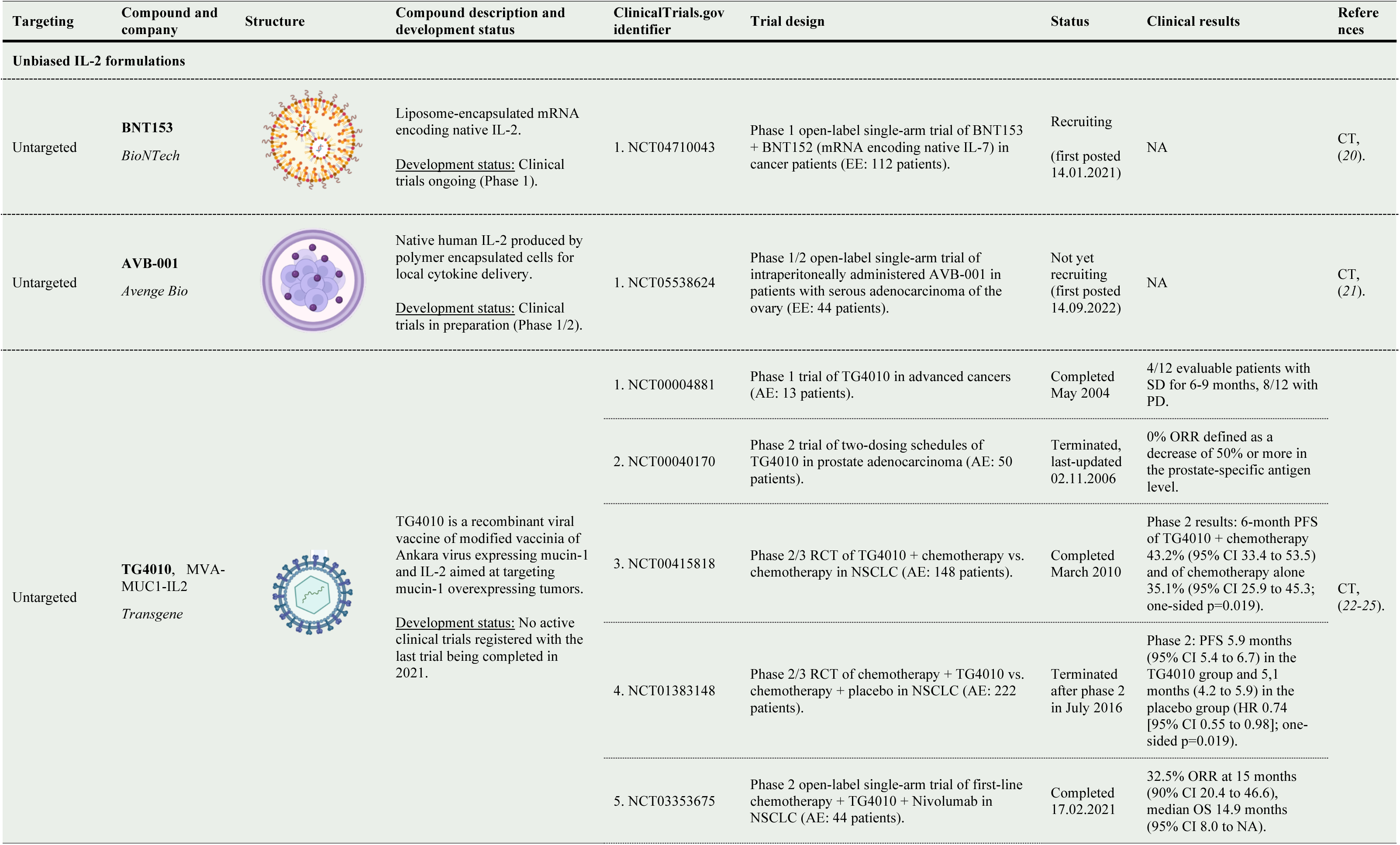
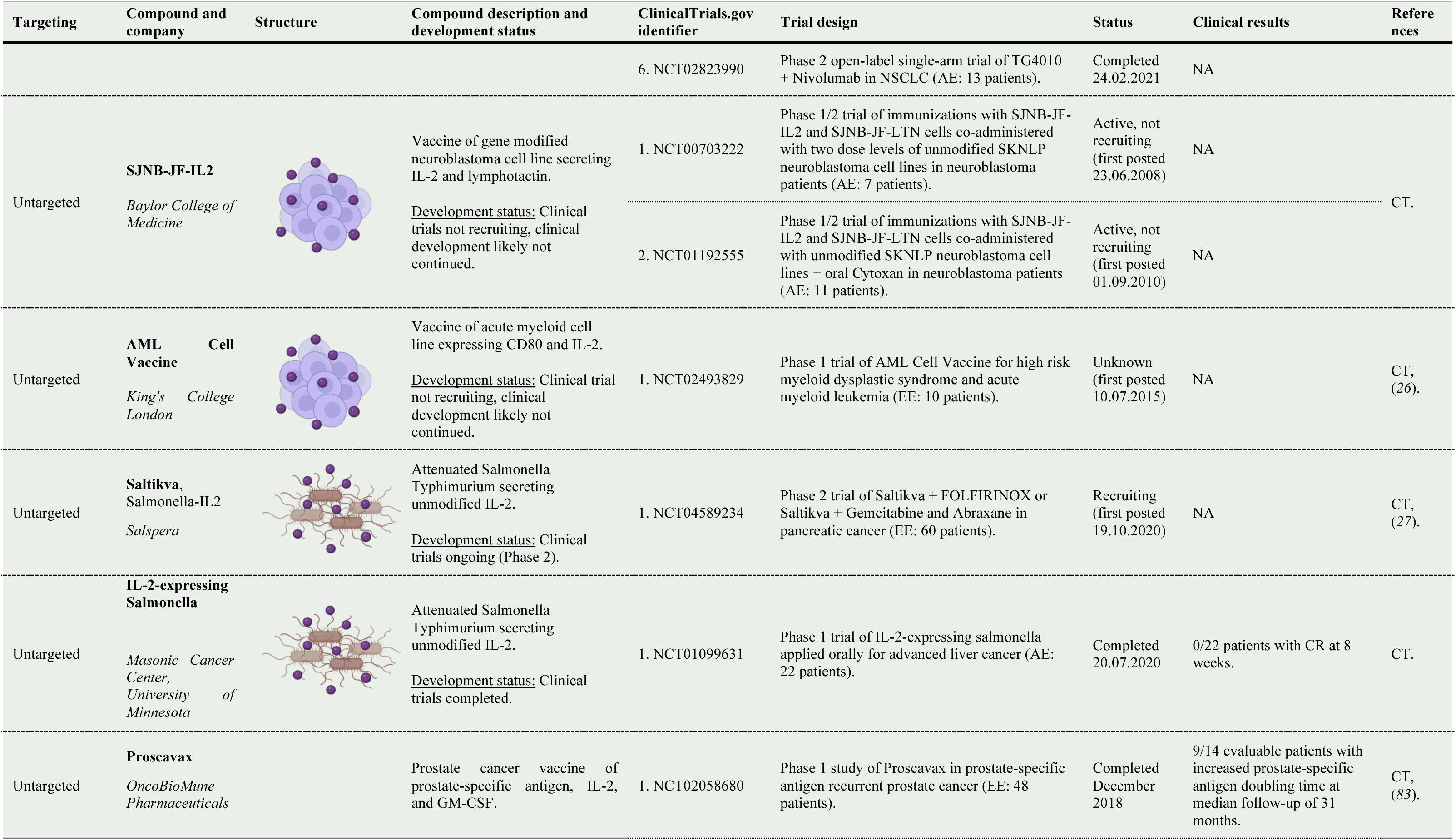
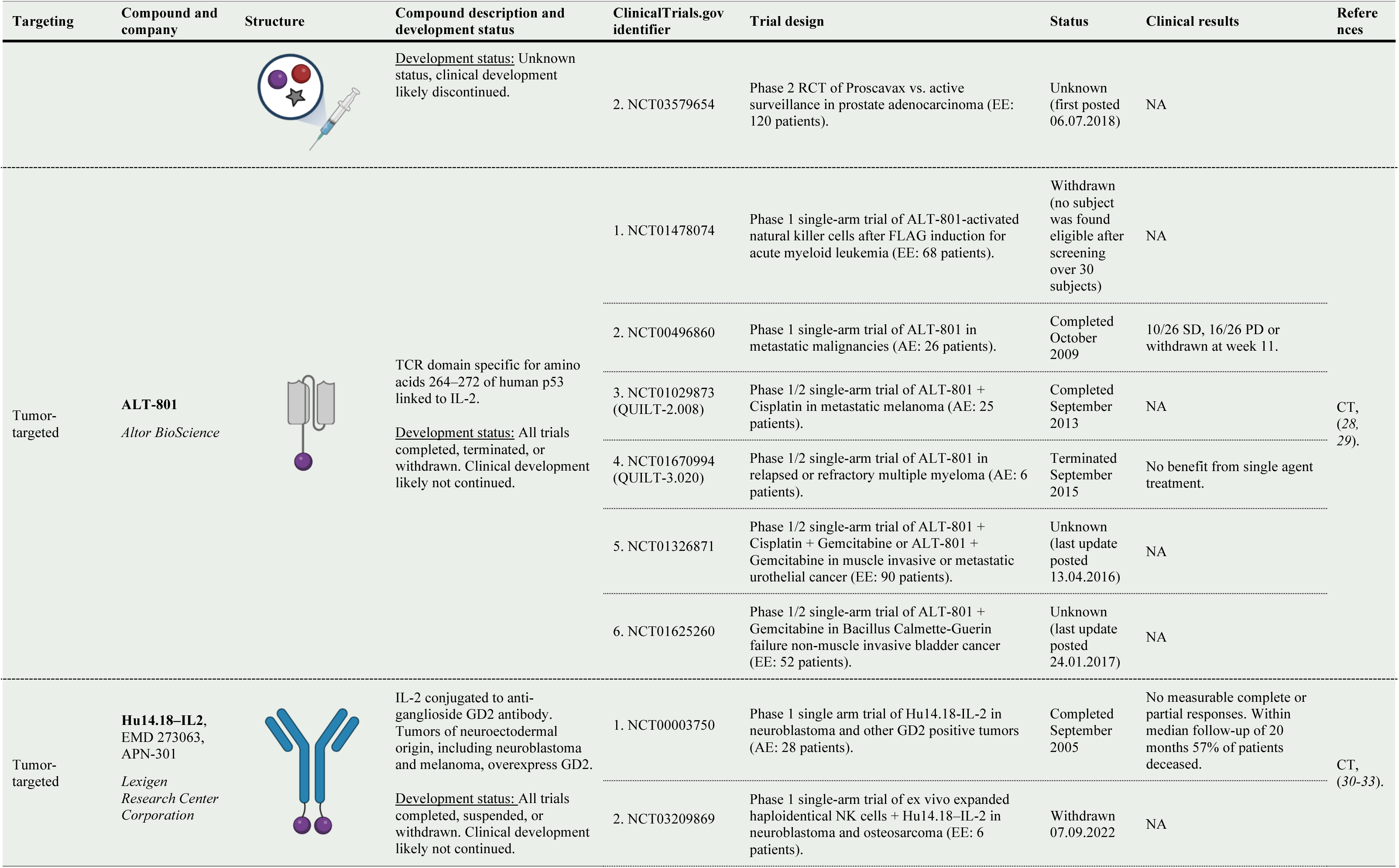
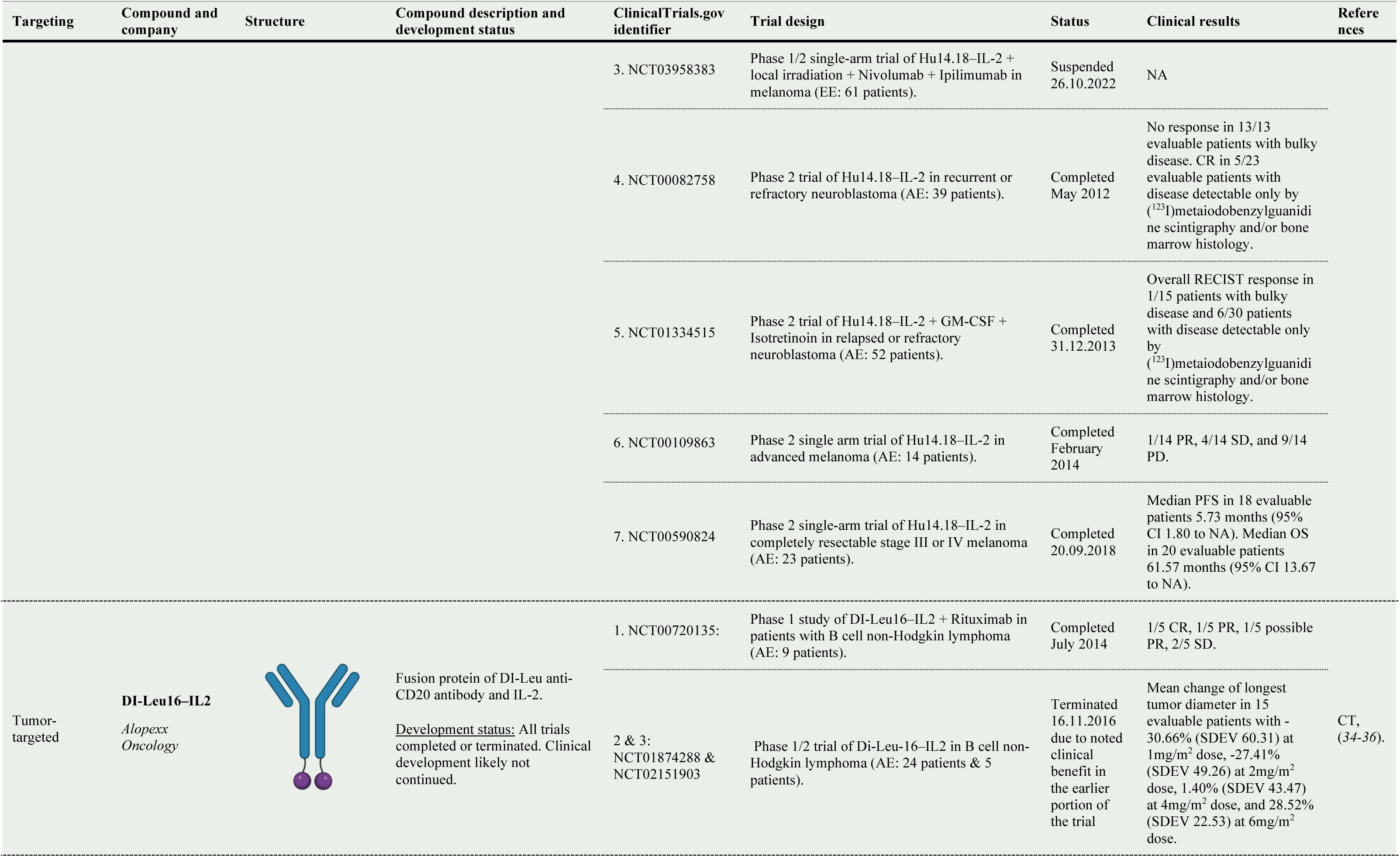
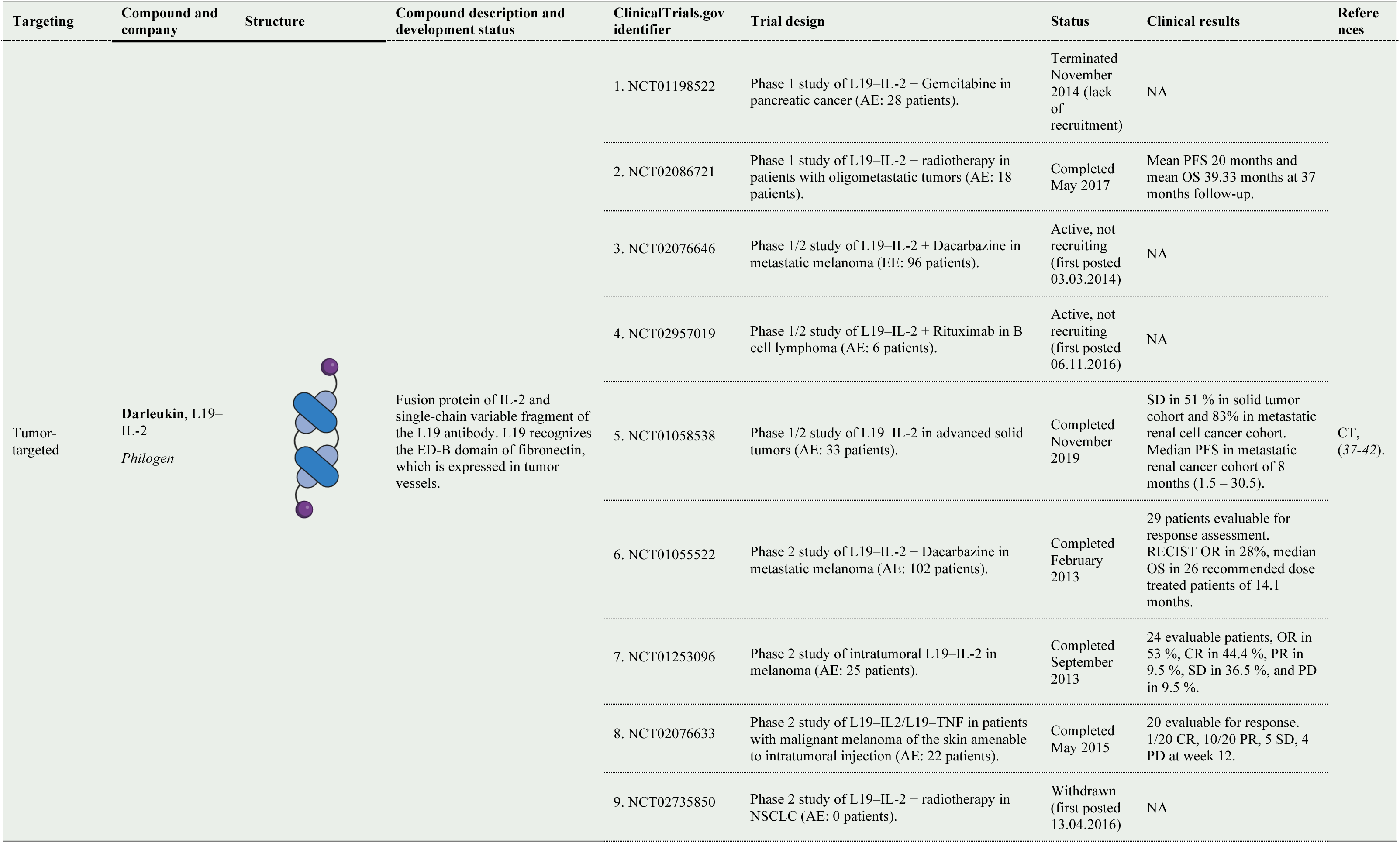
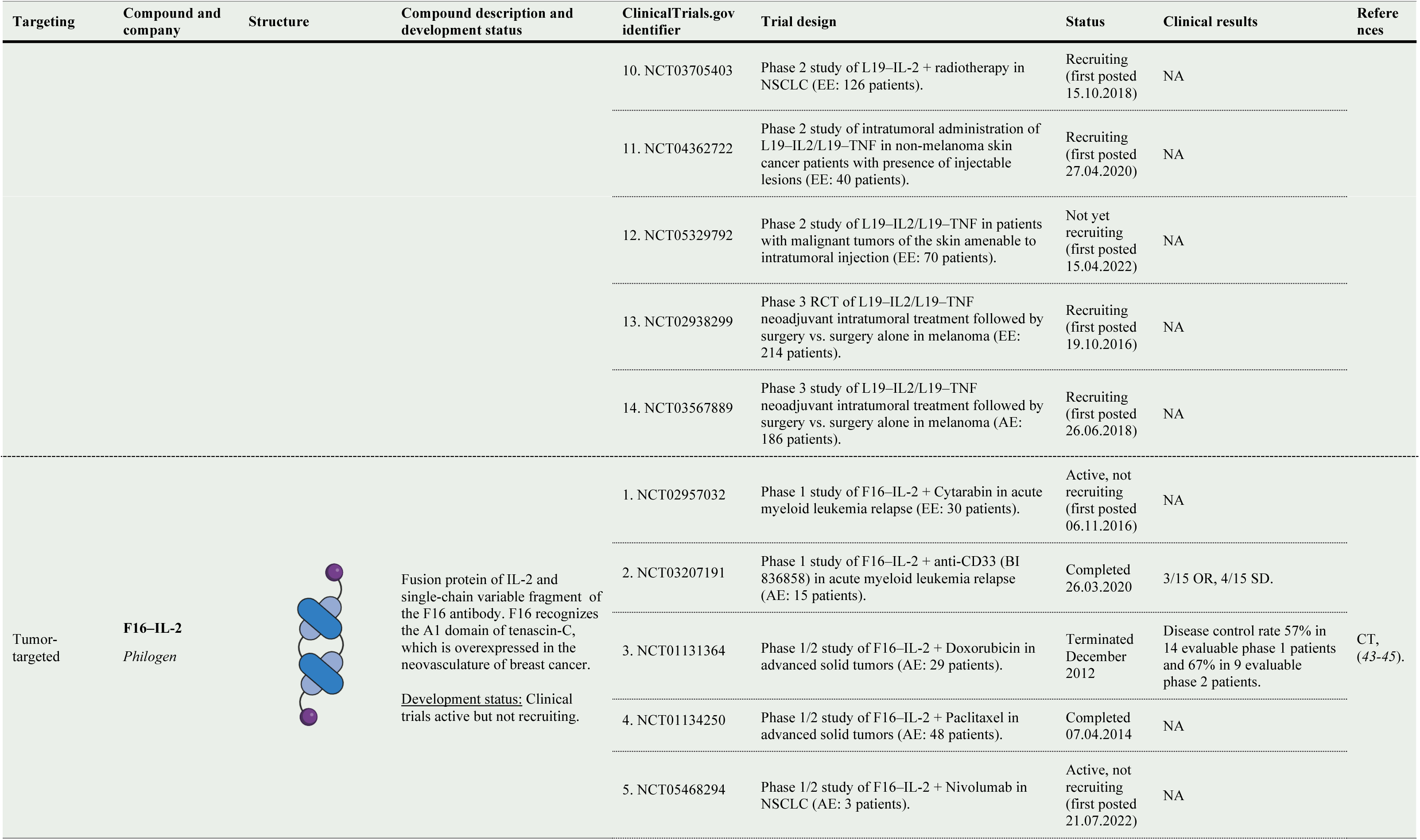
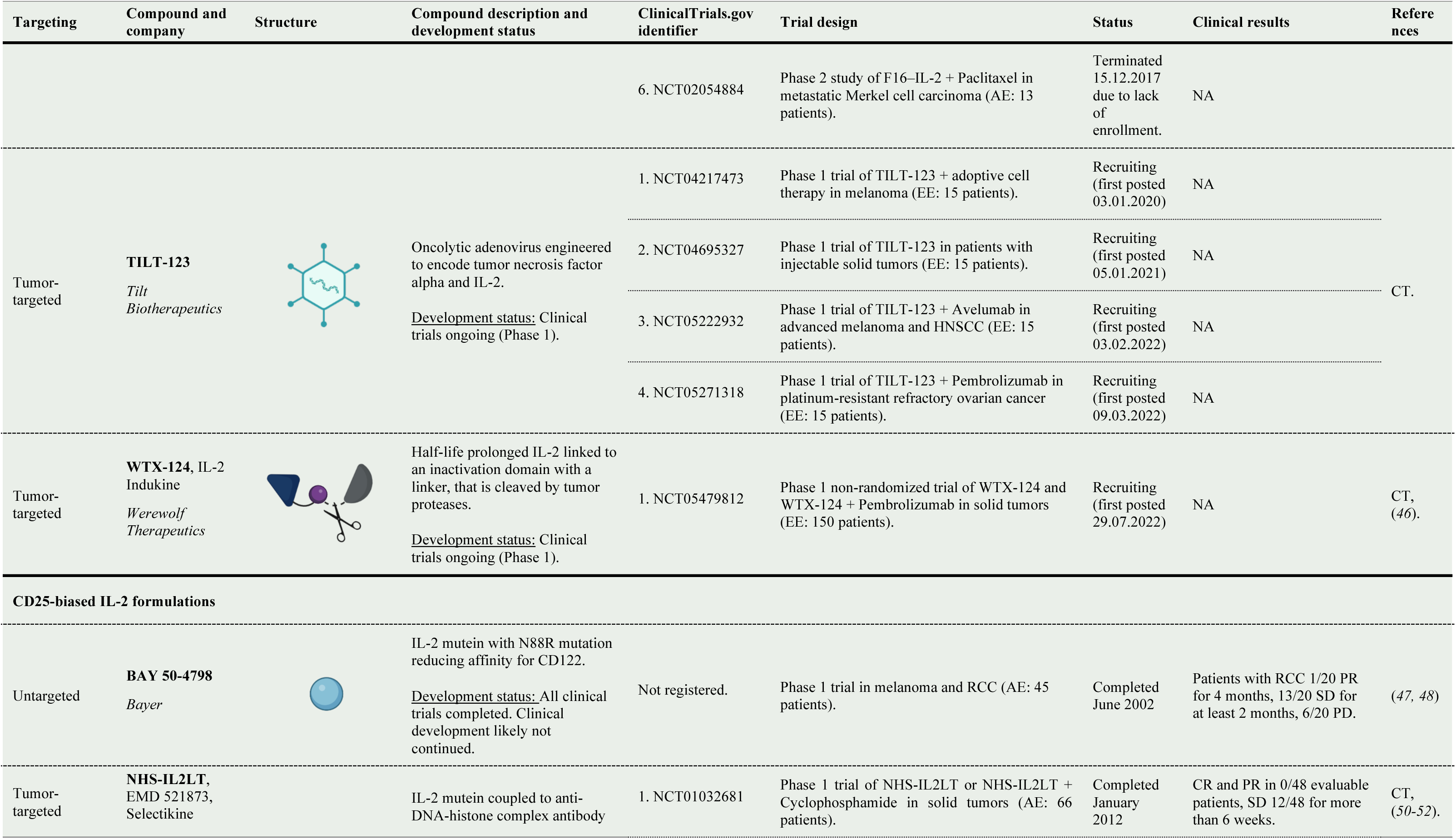
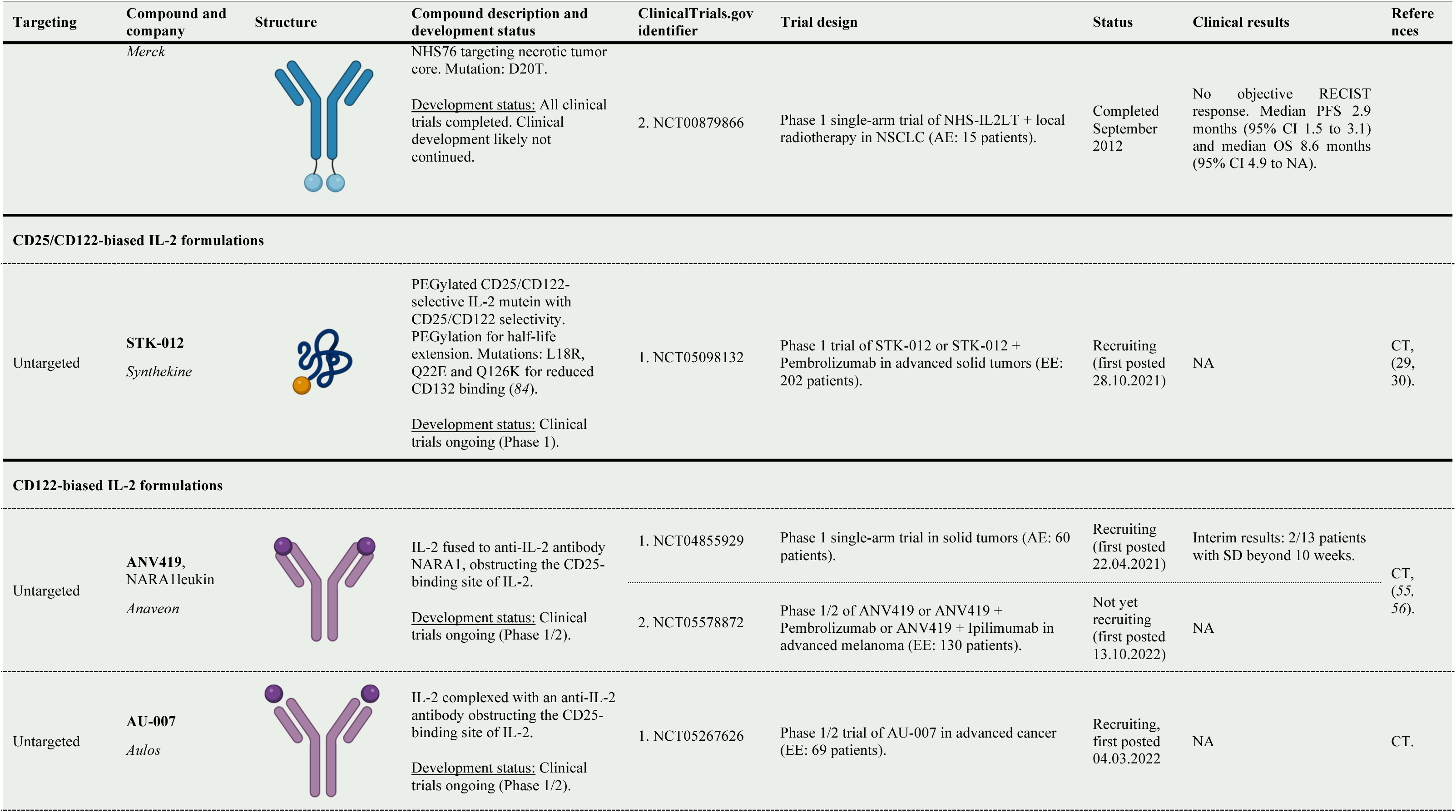
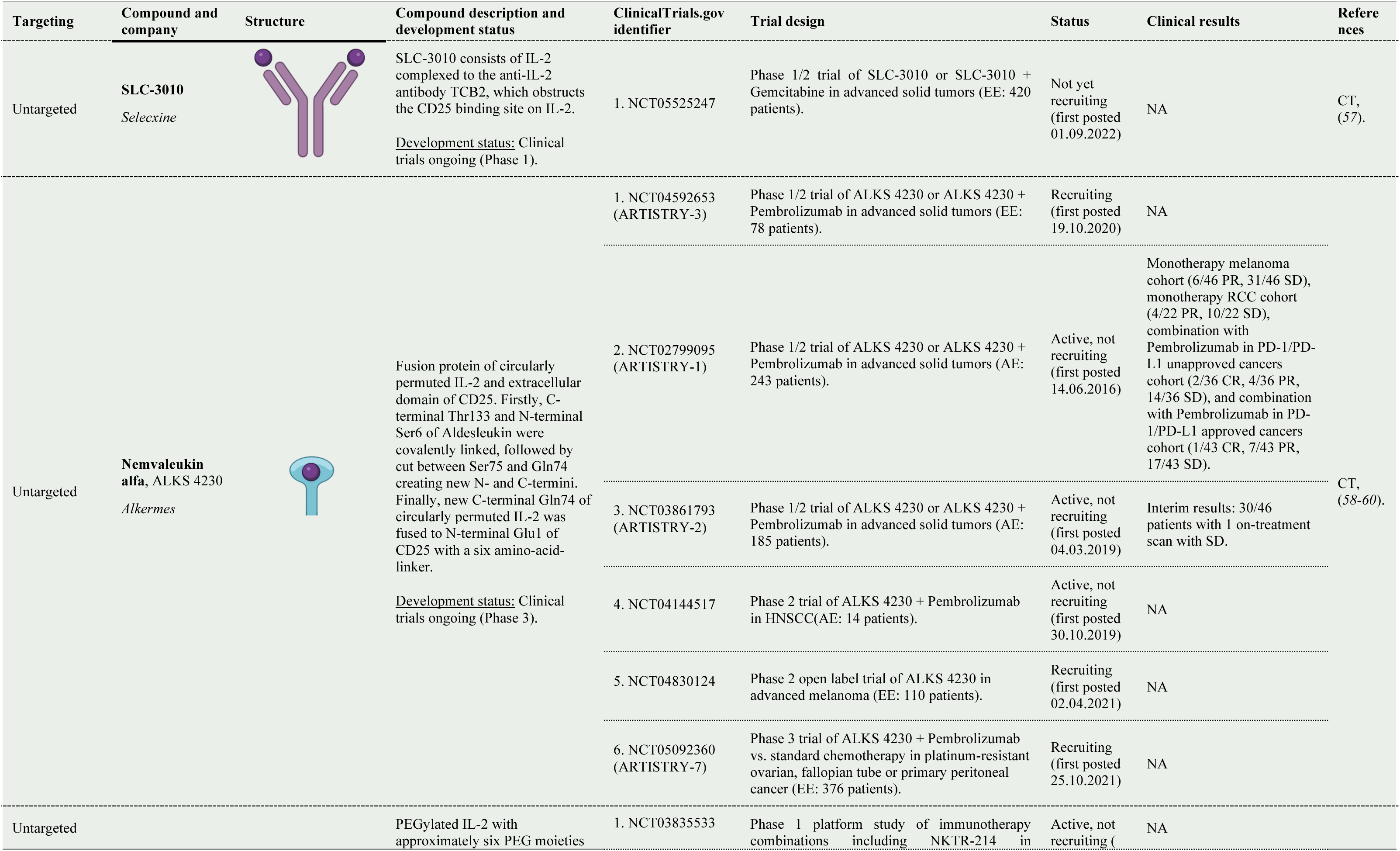
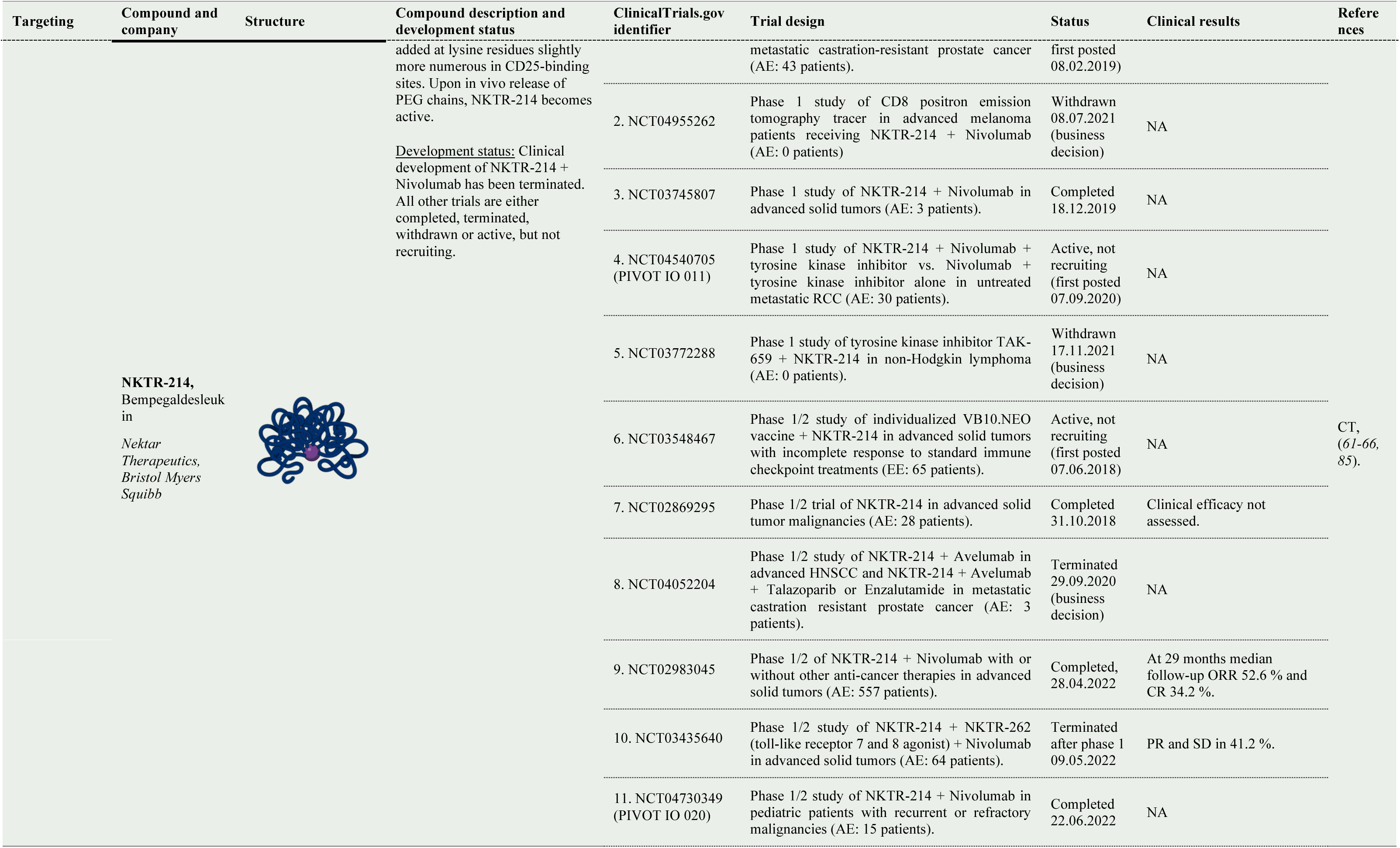
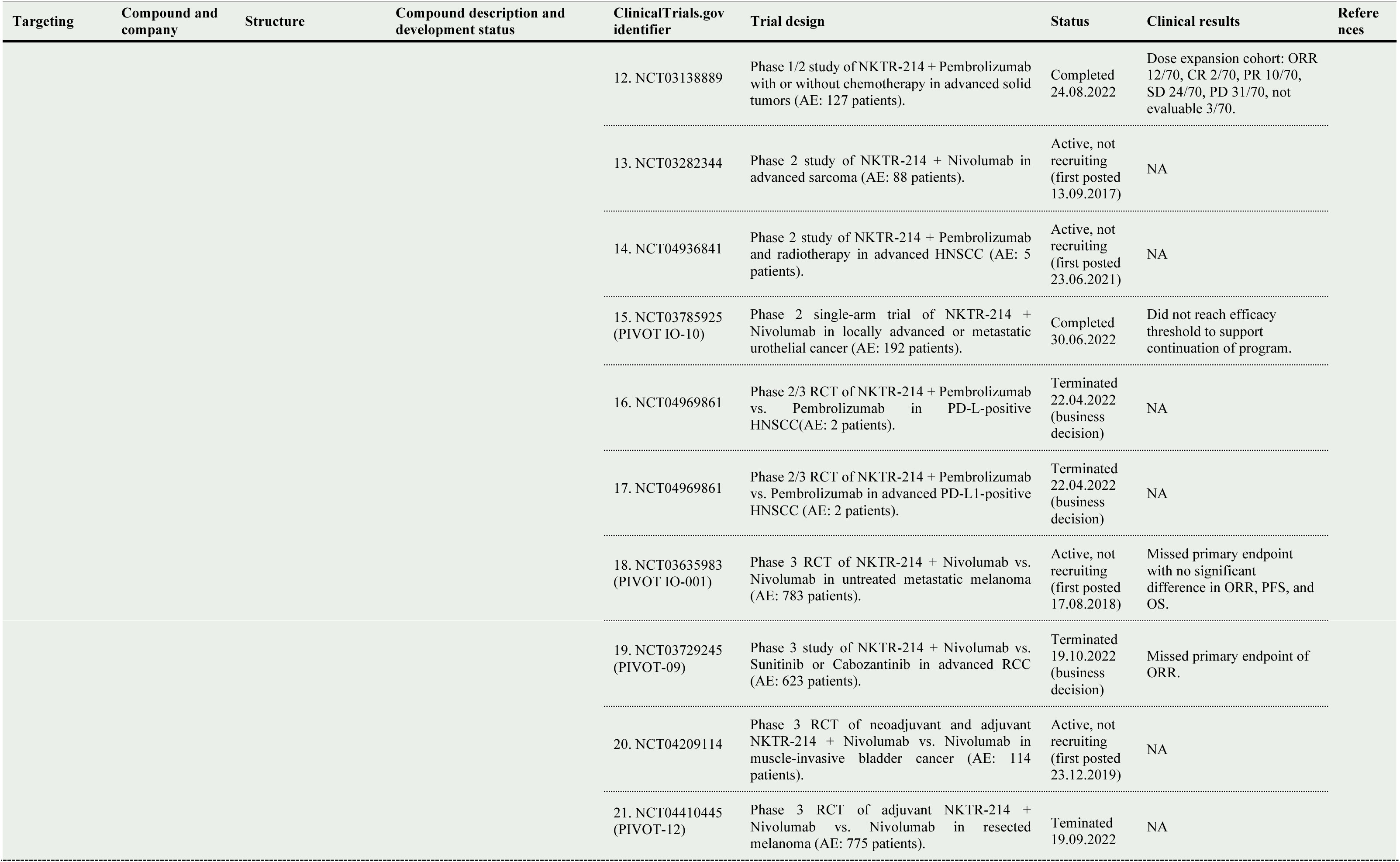
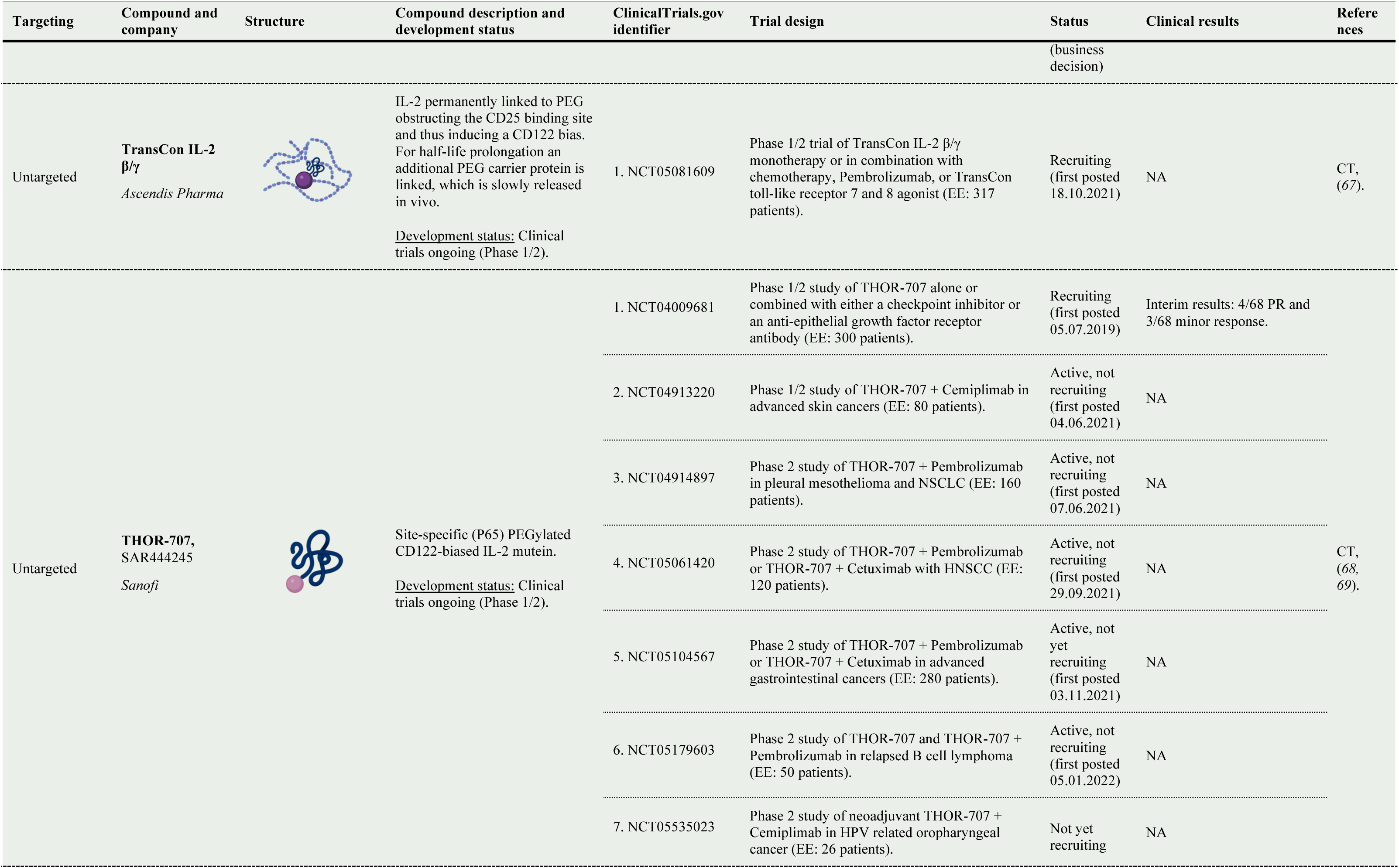
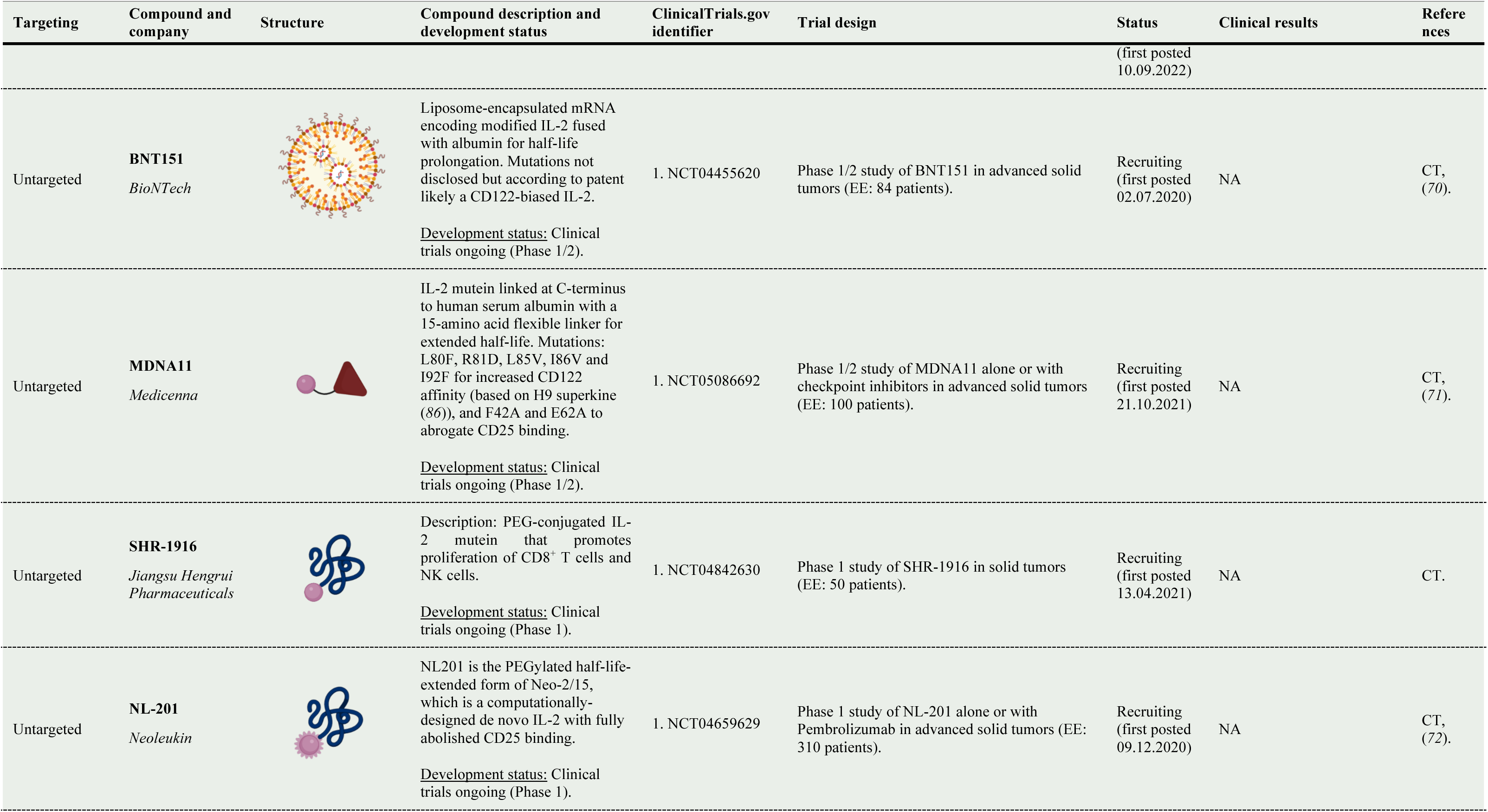
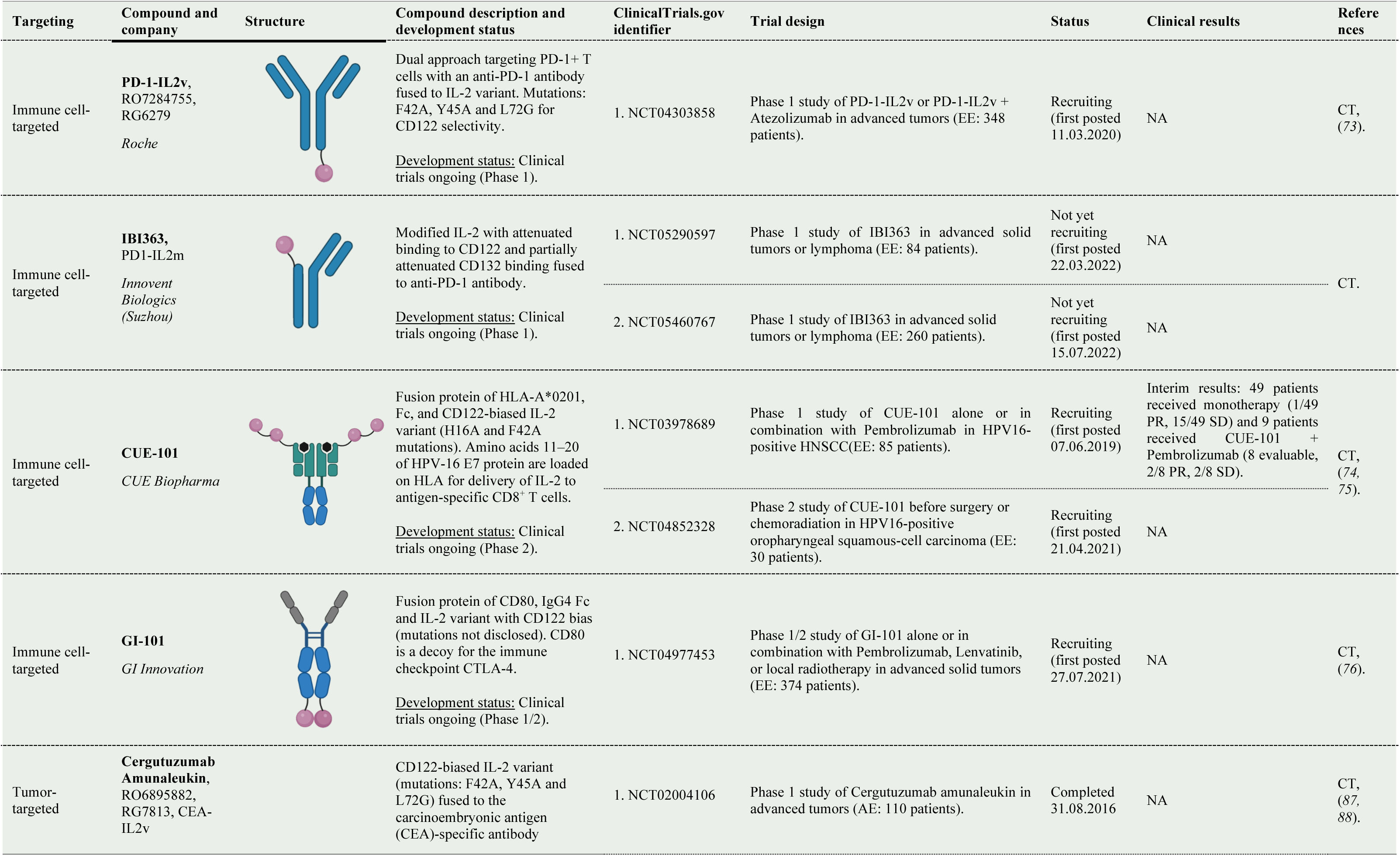
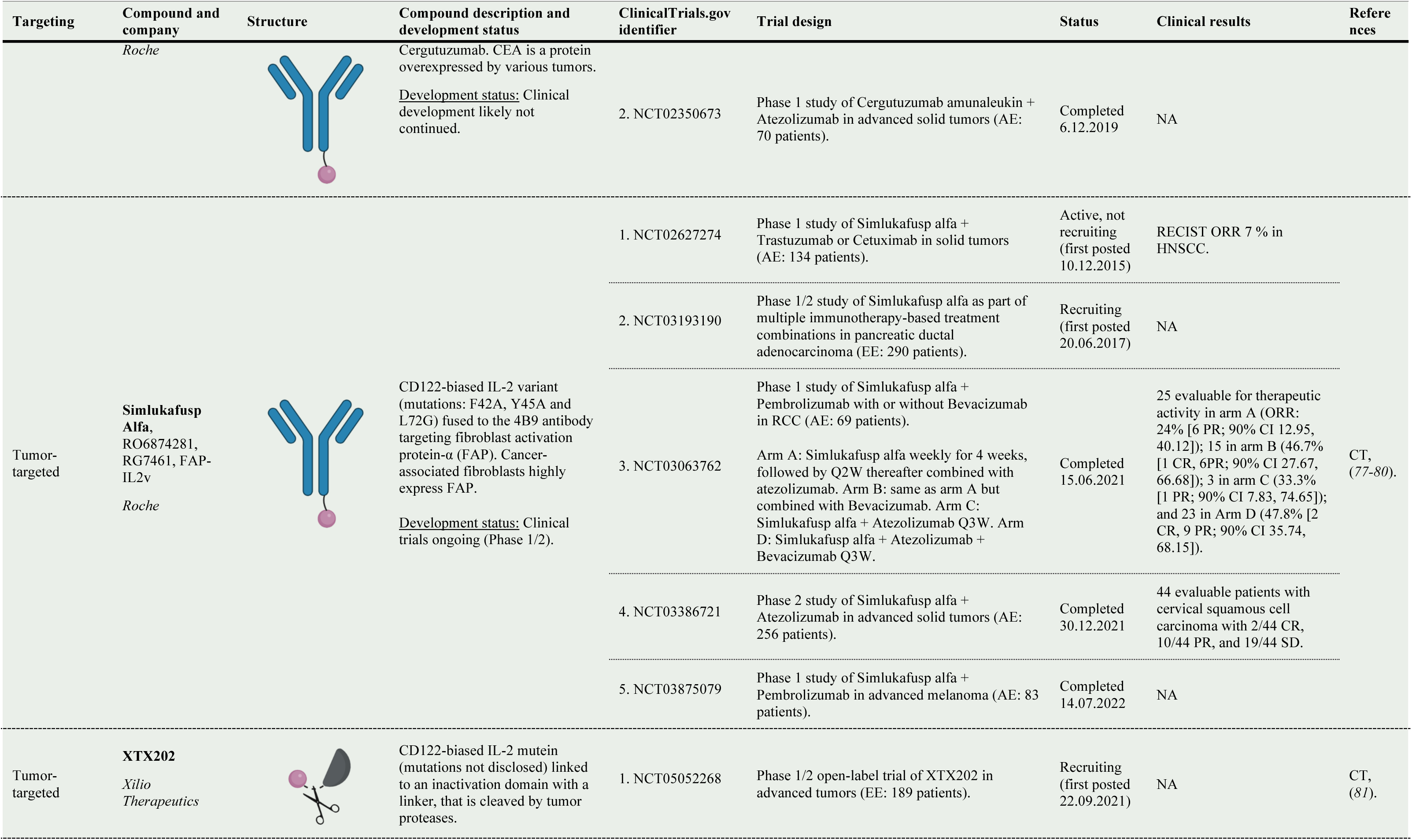
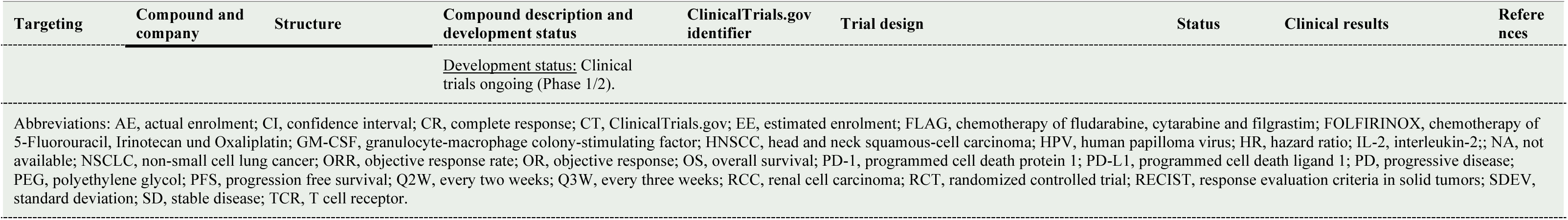
Improved IL-2-based compounds for the treatment of cancer.

**Table 2:**
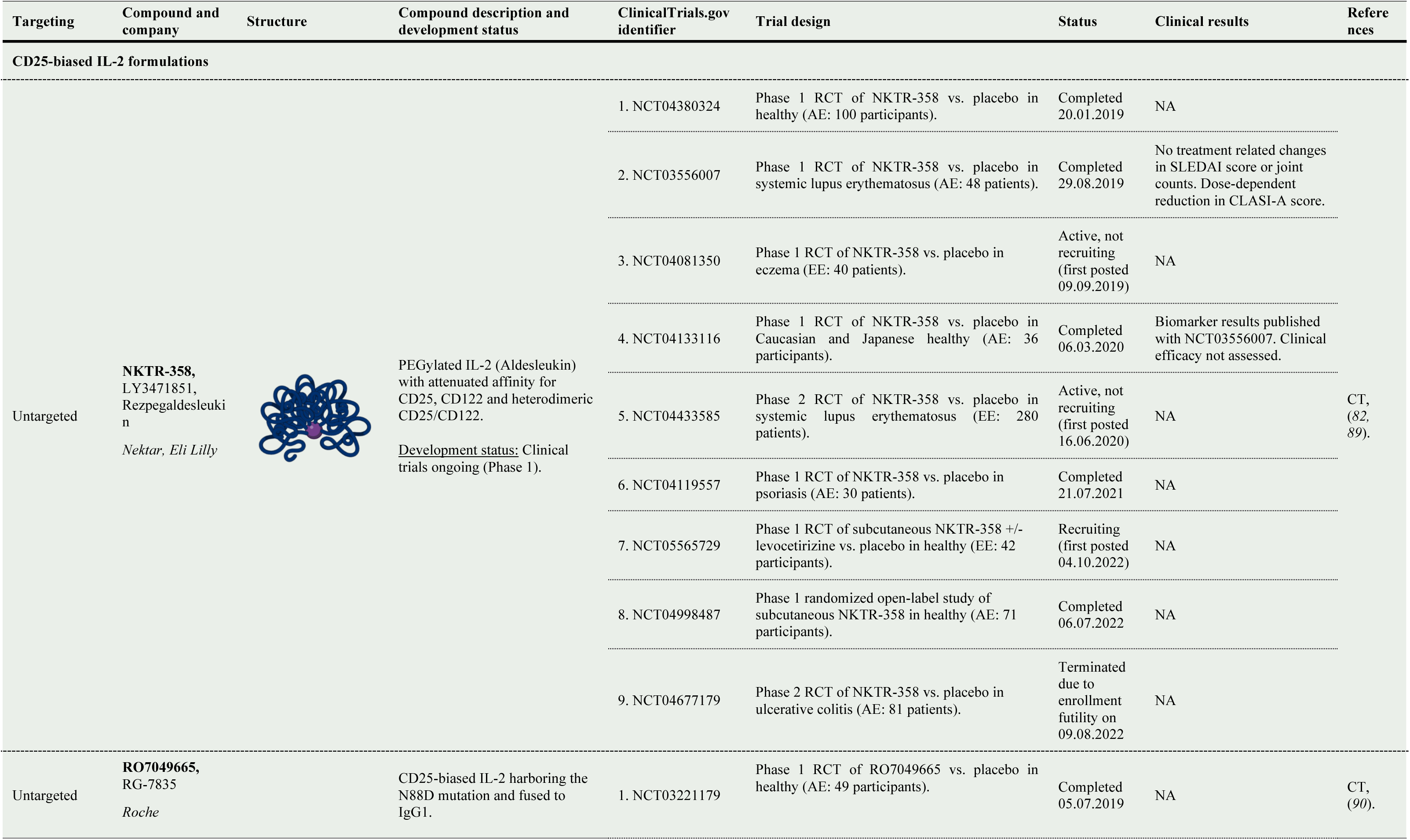
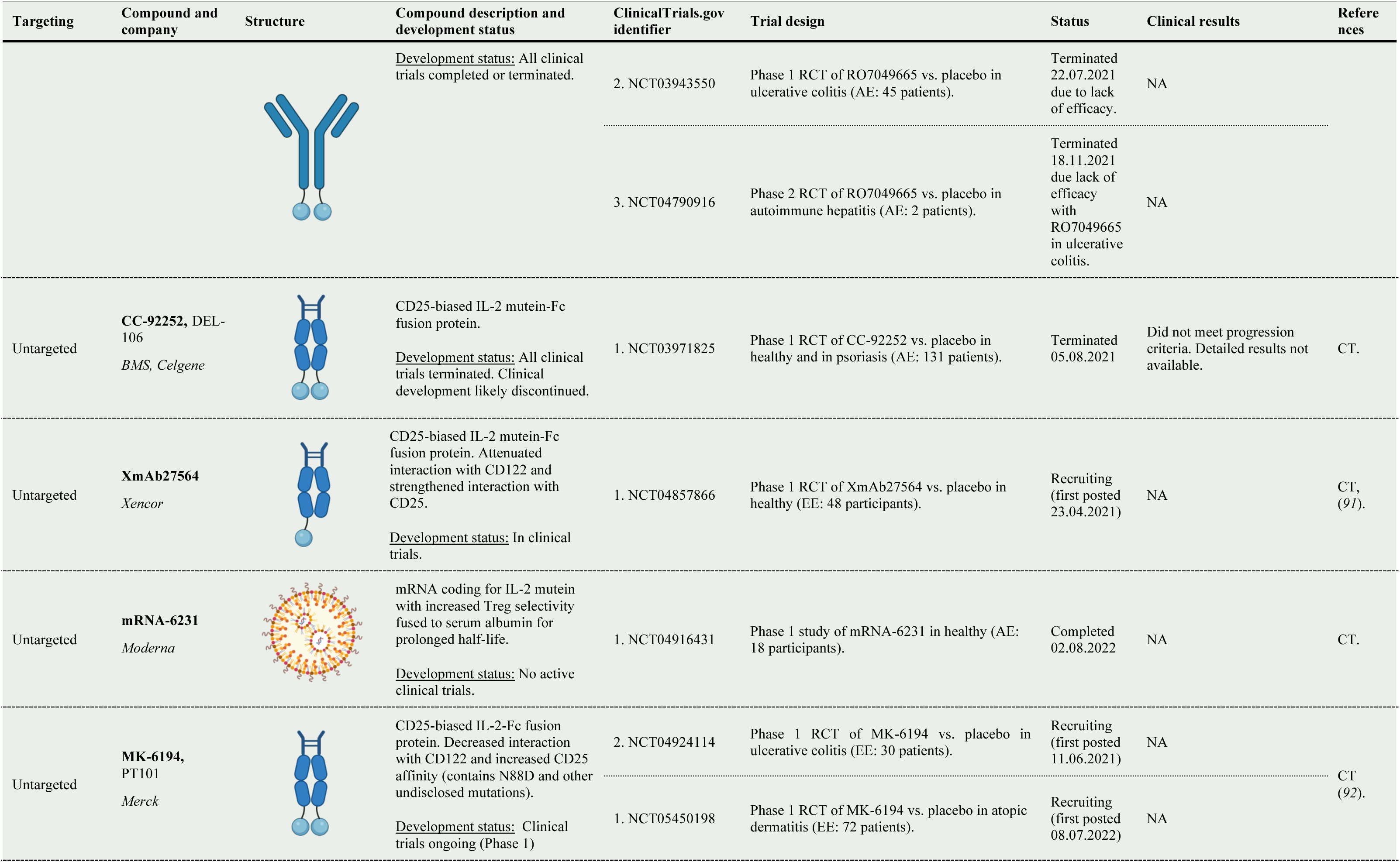
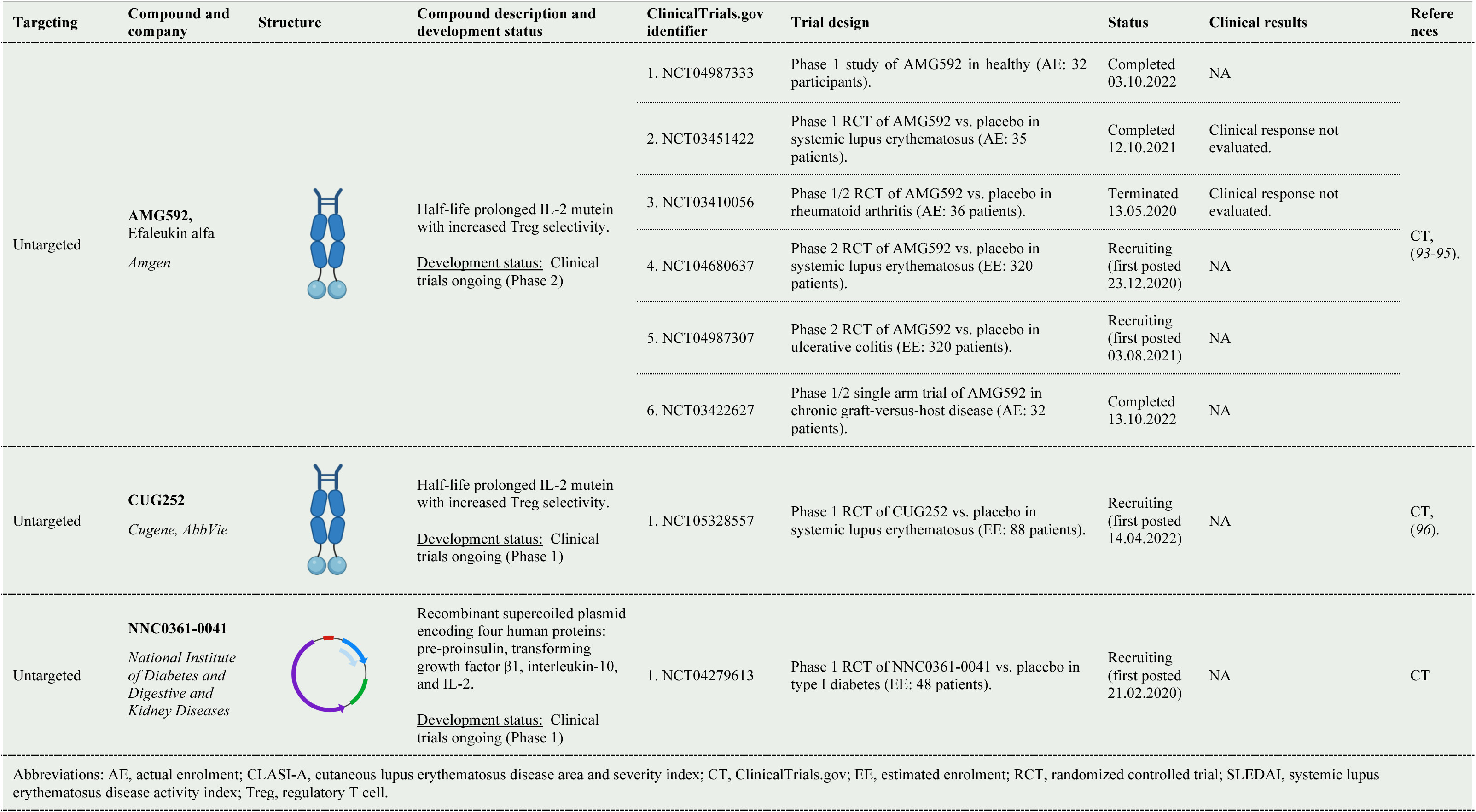
Improved IL-2-based compounds for the treatment of autoimmune diseases.

| Targeting | Compound and company | Structure | Compound description and development status | ClinicalTrials.gov identifier | Trial design | Status | Clinical results | References |
| --- | --- | --- | --- | --- | --- | --- | --- | --- |
| CD25-biased IL-2 formulations |  |  |  |  |  |  |  |  |
| Untargeted | **NKTR-358**,<br>LY3471851,<br>Rezpegaldesleukin<br>*Nektar, Eli Lilly* |  | PEGylated IL-2 (Aldesleukin) with attenuated affinity for CD25, CD122 and heterodimeric CD25/CD122.<br><u>Development status:</u> Clinical trials ongoing (Phase 1). | 1. NCT04380324 | Phase 1 RCT of NKTR-358 vs. placebo in healthy (AE: 100 participants). | Completed 20.01.2019 | NA | CT, (82, 89). |
|  |  |  |  | 2. NCT03556007 | Phase 1 RCT of NKTR-358 vs. placebo in systemic lupus erythematosus (AE: 48 patients). | Completed 29.08.2019 | No treatment related changes in SLEDAI score or joint counts. Dose-dependent reduction in CLASI-A score. |  |
|  |  |  |  | 3. NCT04081350 | Phase 1 RCT of NKTR-358 vs. placebo in eczema (EE: 40 patients). | Active, not recruiting (first posted 09.09.2019) | NA |  |
|  |  |  |  | 4. NCT04133116 | Phase 1 RCT of NKTR-358 vs. placebo in Caucasian and Japanese healthy (AE: 36 participants). | Completed 06.03.2020 | Biomarker results published with NCT03556007. Clinical efficacy not assessed. |  |
|  |  |  |  | 5. NCT04433585 | Phase 2 RCT of NKTR-358 vs. placebo in systemic lupus erythematosus (EE: 280 patients). | Active, not recruiting (first posted 16.06.2020) | NA |  |
|  |  |  |  | 6. NCT04119557 | Phase 1 RCT of NKTR-358 vs. placebo in psoriasis (AE: 30 patients). | Completed 21.07.2021 | NA |  |
|  |  |  |  | 7. NCT05565729 | Phase 1 RCT of subcutaneous NKTR-358 +/- levocetirizine vs. placebo in healthy (EE: 42 participants). | Recruiting (first posted 04.10.2022) | NA |  |
|  |  |  |  | 8. NCT04998487 | Phase 1 randomized open-label study of subcutaneous NKTR-358 in healthy (AE: 71 participants). | Completed 06.07.2022 | NA |  |
|  |  |  |  | 9. NCT04677179 | Phase 2 RCT of NKTR-358 vs. placebo in ulcerative colitis (AE: 81 patients). | Terminated due to enrollment futility on 09.08.2022 | NA |  |
| Untargeted | **RO7049665**,<br>RG-7835<br>*Roche* |  | CD25-biased IL-2 harboring the N88D mutation and fused to IgG1. | 1. NCT03221179 | Phase 1 RCT of RO7049665 vs. placebo in healthy (AE: 49 participants). | Completed 05.07.2019 | NA | CT, (90). |
|  |  |  | <u>Development status:</u> All clinical trials completed or terminated. | 2. NCT03943550 | Phase 1 RCT of RO7049665 vs. placebo in ulcerative colitis (AE: 45 patients). | Terminated 22.07.2021 due to lack of efficacy. | NA |  |
|  |  |  |  | 3. NCT04790916 | Phase 2 RCT of RO7049665 vs. placebo in autoimmune hepatitis (AE: 2 patients). | Terminated 18.11.2021 due lack of efficacy with RO7049665 in ulcerative colitis. | NA |  |
| Untargeted | **CC-92252**, DEL-106<br>*BMS, Celgene* |  | CD25-biased IL-2 mutein-Fc fusion protein.<br><u>Development status:</u> All clinical trials terminated. Clinical development likely discontinued. | 1. NCT03971825 | Phase 1 RCT of CC-92252 vs. placebo in healthy and in psoriasis (AE: 131 patients). | Terminated 05.08.2021 | Did not meet progression criteria. Detailed results not available. | CT. |
| Untargeted | **XmAb27564**<br>*Xencor* |  | CD25-biased IL-2 mutein-Fc fusion protein. Attenuated interaction with CD122 and strengthened interaction with CD25.<br><u>Development status:</u> In clinical trials. | 1. NCT04857866 | Phase 1 RCT of XmAb27564 vs. placebo in healthy (EE: 48 participants). | Recruiting (first posted 23.04.2021) | NA | CT, (91). |
| Untargeted | **mRNA-6231**<br>*Moderna* |  | mRNA coding for IL-2 mutein with increased Treg selectivity fused to serum albumin for prolonged half-life.<br><u>Development status:</u> No active clinical trials. | 1. NCT04916431 | Phase 1 study of mRNA-6231 in healthy (AE: 18 participants). | Completed 02.08.2022 | NA | CT. |
| Untargeted | **MK-6194**, PT101<br>*Merck* |  | CD25-biased IL-2-Fc fusion protein. Decreased interaction with CD122 and increased CD25 affinity (contains N88D and other undisclosed mutations).<br><u>Development status:</u> Clinical trials ongoing (Phase 1) | 2. NCT04924114<br>1. NCT05450198 | Phase 1 RCT of MK-6194 vs. placebo in ulcerative colitis (EE: 30 patients).<br>Phase 1 RCT of MK-6194 vs. placebo in atopic dermatitis (EE: 72 patients). | Recruiting (first posted 11.06.2021)<br>Recruiting (first posted 08.07.2022) | NA<br>NA | CT (92). |
| Untargeted | **AMG592**,<br>Efaleukin alfa<br>*Amgen* |  | Half-life prolonged IL-2 mutein with increased Treg selectivity.<br><u>Development status:</u> Clinical trials ongoing (Phase 2) | 1. NCT04987333 | Phase 1 study of AMG592 in healthy (AE: 32 participants). | Completed<br>03.10.2022 | NA | CT,<br>(93-95). |
|  |  |  |  | 2. NCT03451422 | Phase 1 RCT of AMG592 vs. placebo in systemic lupus erythematosus (AE: 35 patients). | Completed<br>12.10.2021 | Clinical response not evaluated. |  |
|  |  |  |  | 3. NCT03410056 | Phase 1/2 RCT of AMG592 vs. placebo in rheumatoid arthritis (AE: 36 patients). | Terminated<br>13.05.2020 | Clinical response not evaluated. |  |
|  |  |  |  | 4. NCT04680637 | Phase 2 RCT of AMG592 vs. placebo in systemic lupus erythematosus (EE: 320 patients). | Recruiting<br>(first posted 23.12.2020) | NA |  |
|  |  |  |  | 5. NCT04987307 | Phase 2 RCT of AMG592 vs. placebo in ulcerative colitis (EE: 320 patients). | Recruiting<br>(first posted 03.08.2021) | NA |  |
|  |  |  |  | 6. NCT03422627 | Phase 1/2 single arm trial of AMG592 in chronic graft-versus-host disease (AE: 32 patients). | Completed<br>13.10.2022 | NA |  |
| Untargeted | **CUG252**<br>*Cugene, AbbVie* |  | Half-life prolonged IL-2 mutein with increased Treg selectivity.<br><u>Development status:</u> Clinical trials ongoing (Phase 1) | 1. NCT05328557 | Phase 1 RCT of CUG252 vs. placebo in systemic lupus erythematosus (EE: 88 patients). | Recruiting<br>(first posted 14.04.2022) | NA | CT,<br>(96). |
| Untargeted | **NNC0361-0041**<br>*National Institute of Diabetes and Digestive and Kidney Diseases* | | Recombinant supercoiled plasmid encoding four human proteins: pre-proinsulin, transforming growth factor β1, interleukin-10, and IL-2.<br><u>Development status:</u> Clinical trials ongoing (Phase 1) | 1. NCT04279613 | Phase 1 RCT of NNC0361-0041 vs. placebo in type I diabetes (EE: 48 patients). | Recruiting<br>(first posted 21.02.2020) | NA | CT |
| Abbreviations: AE, actual enrolment; CLASI-A, cutaneous lupus erythematosus disease area and severity index; CT, ClinicalTrials.gov; EE, estimated enrolment; RCT, randomized controlled trial; SLEDAI, systemic lupus erythematosus disease activity index; Treg, regulatory T cell. |  |  |  |  |  |  |  |  |

### Risk of bias assessment

A Modified Downs and Black tool was applied to assess bias of retrieved randomized controlled trials reporting clinical results (**Table S2**) (*19*). Scoring the studies according to the categories (a) reporting, (b) external validity, (c) internal validity and (d) power resulted in ranks of high (23–28 points), medium (15–22 points), and low (0–14 points) quality. We searched the ClinicalTrials.gov database for all studies registered and conducted a secondary search for results available for these trials on MEDLINE, company websites, news sites, and other websites, reducing publication bias.

### Principal summary measures and synthesis of results

We aimed to provide a systematic review of clinical results of improved IL-2-based compounds for the treatment of cancer and autoimmune diseases. To avoid exclusion of IL-2-based compounds, we avoided further specification of these endpoints.

## 3. Results

### Study selection and characteristics

Applying the predefined search terms, we identified and screened 547 clinical trials registered on ClincialTrials.gov. Of these, 511 records were excluded resulting in 36 individual improved IL-2-based compounds. We added another 9 compounds identified by recent literature reviews and based on our knowledge, resulting in a total of 45 compounds. A secondary search for clinical trials of each individual compound resulted in a total of 139 clinical trials that were included in this systematic review, of which 29 had available clinical results.

### Synthesized findings

The findings for improved IL-2-based compounds for the treatment of cancer are synthesized in **Table 1**, and those for the treatment of autoimmune diseases in **Table 2**.

#### Improved IL-2-based compounds in clinical trials for the treatment of cancer

##### Unbiased IL-2-based compounds

The category of unbiased and untargeted IL-2-based compounds comprises several compounds. **BNT153** is a liposome-encapsulated messenger RNA (mRNA) formulation encoding native IL-2 (*20*). Limited published data are available from one phase 1 clinical trial registered testing BNT153 in combination with BNT152, the latter containing mRNA encoding native IL-7, in cancer patients, with no results available as of this date. **AVB-001** is a polymer encapsulated IL-2-producing cellular cytokine factory designed for local delivery (*21*). A phase 1/2 trial of intraperitoneally administered AVB-001 in ovarian serous adenocarcinoma was recently initiated, no results available yet. Another approach is the delivery of IL-2 by viral vectors. **TG4010** is a recombinant vaccine based on modified vaccinia virus Ankara expressing mucin-1 and IL-2, targeting mucin- 1-overexpressing tumors with six registered clinical trials (*22–25*). The phase 2/3 randomized-controlled trial (RCT) NCT02823990 testing standard chemotherapy with TG4010 or placebo enrolling 222 non-small cell lung cancer (NSCLC) patients reported progression-free survival (PFS) of 5.9 months (95% confidence interval [CI] 5.4 to 6.7) in the TG4010 group and 5.1 months (95% CI 4.2 to 5.9) in the placebo group (Hazard ratio [HR] 0,74 [95% CI 0,55 to 0,98]; one-sided p=0.019). The trial was terminated in phase 3 in July 2016.

The latest phase 2 single-arm trial NCT03353675 testing chemotherapy with TG4010 and the immune checkpoint inhibitor Nivolumab in NSCLC reported 32.5% objective response rate (ORR) at 15 months (90% CI 20.4 to 46.6) with median overall survival (OS) of 14.9 months (95% CI 8.0 to not available [NA]). Currently, no active clinical trials are registered using TG4010, with the last trial completed in 2021. **SJNB- JF-IL2** and **AML Cell Vaccine** are vaccines based on genetically modified IL-2-secreting neuroblastoma and acute myeloid leukemia cell lines, respectively (**Table 1**) (*26*). Three clinical trials were registered in 2008, 2010, and 2015, respectively, and are listed active but not recruiting or unknown. Modified *Salmonella typhimurium* bacteria secreting IL-2 are found in **Saltikva**, which is tried in combination with chemotherapy in pancreatic cancer patients within a phase 2 trial registered in October 2010 and currently recruiting (*27*). A very similar compound of **IL-2-expressing Salmonella** was developed at the University of Minnesota and tested in advanced liver cancer, reporting CR in none of the 22 patients enrolled in the trial (**Table 1**). **Proscavax**, a vaccine based on prostate cancer cells secreting prostate-specific antigen, IL-2 and granulocyte-macrophage colony-stimulating factor (GM-CSF) is registered with one clinical trial that was first posted in July 2018 and whose current status is unknown (**Table 1**).

Another category of improved IL-2-based compounds comprises tumor-targeting molecules. **ALT- 801** consists of native IL-2 linked to a single-chain T cell receptor (TCR) domain recognizing amino acids 264–272 of human p53 antigen, thus shuttling IL-2 to the tumor (*28, 29*). Six clinical trials are registered with all of them either completed, terminated, or withdrawn, indicating that the clinical development has been suspended. Results are available for the NCT00496860 single-arm phase 1 trial testing ALT-801 in 26 patients with metastatic malignancies reporting stable disease (SD) in 10 of 26, and progressive disease (PD) or withdrawn in 16/26 patients. The IL-2 fusion protein **Hu14.18-IL2** consists of two IL-2 molecules fused to an anti-ganglioside GD2 antibody (*30–33*). GD2 is expressed in tumors of neuroectodermal origin, including neuroblastoma and melanoma. Of the seven registered clinical trials all are either completed, suspended, or withdrawn. The most advanced single-arm phase 2 trial testing Hu14.18-IL2 as adjuvant treatment in completely resectable stage III or IV melanoma patients reported PFS in 18 evaluable patients of 5.73 months (95% CI 1.80 to NA) and a median OS in 20 evaluable patients of 61.6 months (95% CI 13.7 to NA). Another fusion protein of IL-2 and a CD20-targeting antibody, **DI-Leu16–IL2**, was tested in a phase 1/2 trial for the treatment of B cell non-Hodgkin lymphoma (*34–36*). The trial NCT 02151903 reported reduction in tumor diameters in 15 evaluable patients of up to 30.7% (standard deviation [SDEV] 60.3) at 1 mg/m^2^ dose. This trial was completed in November 2016 and no additional trials are registered. An alternative strategy used fusion proteins of IL-2 molecules linked to antibody light chains either targeting the ED-B domain of an angiogenesis-associated isoform of fibronectin overexpressed in tumor vessels (**Darleukin**) (*37–42*) or the alternatively spliced A1 domain of tenascin-C strongly expressed in neovascular stroma of breast cancer (**F16– IL-2**) (*43, 44*). **Darleukin** or L19–IL-2 lists 14 registered phase 1 to 3 clinical trials with four reporting clinical results. The phase 1 trial NCT02086721 observed mean PFS of 20 months and mean OS of 39.3 months at 37 months follow-up in patients with oligometastatic tumors. Results of the phase 1/2 study NCT01058538 in advanced solid tumors showed SD in 51% in the solid tumor cohort and 83% in the metastatic RCC cohort, with median PFS in the metastatic RCC cohort of 8 months (1.5–30.5 months). The more advanced phase 2 trial NCT01253096 testing intratumoral injection of Darleukin in melanoma patients reported in 24 evaluable patients OR in 53%, CR in 44.4%, PR in 9.5%, SD in 36.5%, and PD in 9.5%, and the phase 2 study NCT01055522 of Darleukin + Dacarbazine in metastatic melanoma reported in 29 patients evaluable for response assessment RECIST OR in 28% and median OS of 14.1 months in 26 patients treated with the recommended dose (*37–41*). **F16-IL-2** has six registered clinical trials with the phase 1/2 study NCT01131364 testing F16-IL-2 with doxorubicin in advanced solid tumors reporting disease control rate of 57% in 14 evaluable patients. Several clinical trials are listed active but are currently not recruiting (*43–45*). **TILT-123** is an oncolytic adenovirus encoding tumor necrosis factor (TNF) and IL-2, delivering these cytokines directly to the tumor microenvironment. Four phase 1 clinical trials are registered and currently recruiting patients (**Table 1**). Coupling half-life prolonged IL-2 with a linker to an inactivation domain that is cleaved by tumor proteases in the tumor microenvironment (**WTX-124)** allows delivery of IL-2 specifically to the tumor (*46*). One clinical trial of WTX-124 alone or in combination with pembrolizumab is ongoing and recruiting.

##### CD25-biased IL-2 compounds

Due to the adverse effects of high-dose native IL-2, which initially was believed to be due to excessive activation of CD122-expressing NK cells, the first improved IL-2 compound developed was a CD25-biased IL-2 mutein harboring the N88R mutation (*47*). **BAY 50-4798** was tested in a phase 1 trial in patients with RCC reporting PR in 1 of 20 patients (5%), SD in 65% for at least 2 months, and PD in 30% (*48*). Due to limited efficacy and the emergence of neutralizing anti-drug antibodies directed against the mutated sequence of IL-2 further clinical development of BAY 50-4798 was stopped (*49*).

Another CD25-biased IL-2 fusion protein, termed **NHS-IL2LT**, consists of an anti-DNA-histone complex antibody coupled to an IL-2 mutein for targeting the necrotic tumor core. NHS-IL2LT was tested in two phase 1 trials. NCT00879866 testing NHS-IL2LT with radiotherapy in NSCLC reported no objective RECIST response, and NCT01032681 testing NHS-IL2LT either alone or combined with cyclophosphamide observed only SD in 12 of 48 patients with no CR or PR (*50–52*). All registered clinical trials have been completed.

##### CD25/CD122-biased IL-2 compounds

**STK-012** is a pegylated IL-2 mutein with suggested CD25/CD122 selectivity due to mutations reducing binding to CD132 (29, 30). One phase 1 trial of STK-012 alone compared to STK-012 combined with Pembrolizumab in advanced solid tumors is ongoing with no results available yet.

##### CD122-biased IL-2 compounds

The largest category of improved IL-2-based compounds comprises CD122-biased IL-2. The first such compounds developed were so called IL-2–anti-IL-2 mAb complexes, or briefly referred to as IL-2 complexes (*53*). **ANV419** is a second-generation IL-2 complex fusion protein of IL-2 fused to the anti-IL-2 mAb NARA1, the latter of which covers with high affinity the CD25-binding site, thus obstructing association with CD25 (*54, 55*). Interim results of the NCT04855929 phase 1 trial showed SD beyond 10 weeks in 2 of 13 patients (*56*). Two first-generation IL-2 complexes, **AU-007** and **SLC-3010**, where IL-2 is non-covalently complexed to specific anti-IL-2 mAb are also in clinical development. AU-007 and SLC-3010 have each registered one phase 1/2 study, currently recruiting patients (**Table 1**) (*57*).

**Nemvaleukin alfa** is a fusion protein of IL-2 fused to the extracellular domain of CD25, thus obstructing the CD25-binding site and inducing a CD122 bias (*58–60*). Six clinical trials from phase 1 to 3 are registered. Available results of the ARTISTRY-1 trial (NCT02799095) show four of 22 melanoma patients with PR in the monotherapy cohort, two of 36 CR and four of 36 PR in combination with Pembrolizumab in the PD-1/PD-L1-unapproved cancers cohort, and one of 43 CR and seven of 43 PR in combination with Pembrolizumab in the PD-1/PD-L1-approved cancer cohort. A phase 3 trial testing Nemvaleukin alfa combined with Pembrolizumab against standard chemotherapy in platinum-resistant ovarian, fallopian tube or primary peritoneal cancer was initiated in October 2021.

Another approach to induce CD122-bias is achieved by adding polyethylene glycol (PEG) moieties to CD25-binding sites on IL-2, a process called PEGylation. Apart from inducing IL-2R bias, PEGylation additionally prolongs in vivo half-life of the molecules. The first pegylated IL-2-based compound was **NKTR- 214** (Bempegaldesleukin) (*61–66*). With 21 registered clinical trials this is also the most studied IL-2-based compound, however due to reported lack of efficacy in two phase 3 trials the clinical development program of NKTR-214 was terminated in 2022 (*63, 64*). Detailed clinical results have not been released yet. A similar molecule applying an improved targeted PEGylation approach offers **TransCon IL-2βγ** (*67*). One clinical trial is registered and recruiting patients. Another molecule, **THOR-707**, contains a 30-kDa PEG moiety attached to the P65 site of IL-2, thus obstructing CD25 binding (*68, 69*). Six clinical trials are registered, all of them not yet recruiting patients. According to the recently released 2022 third quarter results by Sanofi, the phase 2 platform of THOR-707 was closed for dose optimization within a phase 1 trial (*36*).

Very commonly, specific mutations are introduced into IL-2 to either increase binding affinity to CD122 and/or decrease affinity to CD25, resulting in IL-2 muteins. Similar to BNT153, BioNTech developed **BNT151** which is a liposome-encapsulated mRNA formulation encoding for mutated IL-2 with CD122 bias fused to albumin for half-life prolongation (*70*). One phase 1/2 clinical trial is recruiting cancer patients.

**MDNA-11 and SHR-1916**, two other IL-2 muteins with increased CD122-bias fused to serum albumin for half-life extension are both tested in an ongoing phase 1/2 trial in advanced solid tumors, with patients currently being recruited (**Table 1**) (*71*). **NL-201** is a computationally-designed de novo IL-2 with fully abolished CD25 binding linked to a PEG moiety for half-life prolongation (*72*). One phase 1 trial of NL- 201 alone or combined with pembrolizumab in advanced solid tumors is ongoing.

Another group comprises IL-2-based compounds targeting immune cells. Building on the success of checkpoint inhibitors, fusing IL-2 to an anti-PD-1 antibody thus shuttling IL-2 to effector T cells could be promising. Two antibodies of this class, **PD-1-IL2v** and **IBI363**, are currently in clinical development with registered ongoing phase 1 trials. PD-1-IL2v is tested alone or combined with Atezolizumab in advanced tumors (*73*), and **IBI363** in advanced solid tumors and lymphoma (**Table 1**). **CUE-101** is a fusion protein consisting of an IL-2 variant with increased CD122 bias, Fc fragment, and human leukocyte antigen loaded with amino acids 11–20 of human papilloma virus (HPV)-16 E7 (*74, 75*). Of the two registered trials, interim results of the phase 1 trial in HPV-16-positive head and neck squamous cell carcinoma reports PR in 1 of 49 and SD in 15 of 49 patients receiving CUE-101 monotherapy, and PR in 2 of 8 and SD in 2 of 8 patients receiving CUE-101 plus Pembrolizumab. A phase 2 trial is currently recruiting patients. The CD80–IgG4-Fc– IL-2 variant fusion protein **GI-101** uses CD80 as a decoy for CTLA-4 and an IL-2 with improved CD122 bias for increased stimulation of effector T cells (*76*). One phase 1/2 study of GI-101 alone or in combination with Pembrolizumab, Lenvatinib or local radiotherapy is currently recruiting patients.

Two tumor-targeting compounds consisting of an IL-2 variant with increased CD122 bias coupled to either the carcinoembryonic antigen-specific antibody Cergutuzumab (**Cergutuzumab amunaleukin**) or the fibroblast activation protein-α-targeting antibody 4B9 (**Simlukafusp alfa**) have been developed by Roche (*77–80*). For Cergutuzumab amunaleukin two phase 1 trials have been completed in 2016 and 2019, respectively, with results not yet available. For Simlukafusp alfa five trials are registered with results of the phase 2 study NCT03386721 testing Simlukafusp alfa combined with Atezolizumab in advanced solid tumors reporting CR in 2 of 44 and PR in 10 of 44, and SD in 19 of 44 patients. Finally, **XTX202** is a CD122-biased IL-2 mutein linked to an inactivation domain, which becomes cleaved in the tumor microenvironment, thus activating IL- 2 (*81*). One phase 1/2 trial in advanced tumors has been initiated and is currently recruiting patients.

#### IL-2-based compounds in clinical trials for the treatment of autoimmune diseases

##### CD25-biased IL-2 compounds

For the treatment of autoimmune diseases only nine compounds have registered clinical trials, with all of these compounds harboring an IL-2 molecule with improved CD25 bias. **NKTR-358** is a PEGylated IL-2 with attenuated binding to CD25, CD122, and the heterodimer of CD25 and CD122, thus inducing a Treg cell bias. This is claimed due to measured decreased affinity of NKTR-358 for the CD25 and CD122 monomers, and the heterodimer of CD25 and CD122, which disfavors binding to cells expressing the intermediate affinity IL- 2R (*82*). Of the nine registered trials testing NKTR-358 only one phase 1 study (NCT03556007) reports clinical results, which did not reveal any significant treatment-related changes in the validated SLE Disease Activity Index (SLEDAI) or the counts of affected joints in SLE patients. However, a dose-dependent improvement of skin manifestations measured with the Cutaneous Lupus Erythematosus Disease Area and Severity Index (CLASI-A) was observed. **RO704966** is a CD25-biased IL-2 mutein harboring the N88D mutation fused to IgG for prolonged half-life. Three registered clinical trials are all terminated or completed with clinical results not available. The IL-2 mutein-Fc fusion protein **CC-92252** was tested in one phase 1 RCT, which was terminated early as the progression criteria was not met. Detailed results are not available. Another IL-2 mutein-Fc fusion protein named **XmAb27564** has one registered clinical trial currently recruiting participants. **MK-6194,** an IL-2 mutated at N88D and other non-disclosed sites for preserved CD25 but decreased CD122 binding is currently tested in two clinical trials, both recruiting patients. The half-life extended IL-2 mutein (mutations not disclosed) **AMG592** has six registered phase 1 to 2 trials with four of them completed or terminated and two recruiting patients. Results are not available or clinical response was not assessed. **CUG252** is a half-life extended and mutated molecule tested in one phase 1 RCT currently recruiting patients. **mRNA-6231** is an mRNA formulation coding for an IL-2 mutein fused to albumin for prolonged half-life that was tested in a phase 1 study, which was completed but no results are available. The supercoiled plasmid NNC0361-0041 encoding for pre-proinsulin, transforming growth factor β1 (TGF-β1), interleukin-10 (IL-10), and IL-2 is tested within a phase 1 trial in patients with type-1 diabetes mellitus and is currently recruiting patients.

### Risk of bias assessment

Risk of bias of RCTs reporting clinical results was assessed using a modified Downs and Black checklist, reproduced in **Table S2**.

## 4. Discussion

### Summary of main findings

With 36 improved IL-2-based compounds in clinical trials, cancer comprises the main indication of current endeavors brining a more specific IL-2 compound to the clinic. Although many early-phase trials reported promising results, a recent press releases revealed negative results from the two phase 3 studies using NKTR-214. These included PIVOT IO-001 testing NKTR-214 versus NKTR-214 combined with Pembrolizumab in metastatic melanoma, which failed its primary endpoints of ORR, PFS, and OS (*63*). Likewise, the PIVOT-09 study of NKTR-214 combined with Nivolumab compared to Sunitinib or Cabozantinib in advanced renal cell carcinoma missed its primary endpoint of improved ORR. Unfortunately, detailed results have not been made publicly available so far. One could speculate on two reasons for the failure of NKTR-214: Firstly, the targeted indication might be wrong, as melanoma is considered an immunogenic tumor already showing high response rates to checkpoint inhibitor treatment, where NKTR-214 might not provide additional benefit. Accordingly, poorly immunogenic tumors with low response rates to checkpoint inhibitors might be a more suitable target for IL-2-based treatments, as IL-2 might render tumors more immunogenic and thus amenable to checkpoint inhibitor treatment (*8*). Secondly, NKTR-214 constitutes a molecule with a suboptimal design. The PEGylation of approximately six PEG moieties linked to lysine residues, which are slowly released in vivo and are slightly more abundant on the CD25-binding site than on the CD122 epitope of IL-2, might result in an suboptimal CD122 bias or even in a CD25 bias. The future will show if other improved IL-2-based compounds can hold up to their promises and show improved anti-tumor responses, either by featuring a better design, by their selectivity for immune or tumor cells, or by being tried in less immunogenic tumors. The only other compound with available RCT results is TG4010, showing significant efficacy when combined with chemotherapy versus chemotherapy alone in two separate phase 2 studies in NSCLC (NCT01383148 and NCT00415818). As currently no active clinical trials are registered, we suspect that the development of TG4010 was suspended; this could likely be due to the recent success of targeted therapeutics in NSCLC, which have replaced chemotherapy as the standard-of-care.

For the treatment of autoimmune diseases results reporting clinical response are only available for NKTR-358 where a phase 1 trial in SLE did not show improvement in the clinical SLEDAI score, although a dose-dependent improvement in the CLASI-A score focused on cutaneous manifestations has been reported.

## Limitations

To our knowledge, this study comprises the first systematic review summarizing the rapidly expanding field of improved IL-2-based compounds. To minimize bias, we applied standard systematic review techniques and included all registered trials into the synthesis of this review. However, this review has several limitations. By limiting the search to ClinicalTrials.gov, trials registered in other registries might have been missed. Moreover, the majority of the retrieved trials are in early phases with only few reporting phase 3 results. Furthermore, due to the commercial interest many trials did not provide clinical results, potentially inducing a reporting bias. Finally, comparison of results from different trials is complex, as trials were conducted in different disease entities with different outcome measures assessed at different time points and different dosing schemes and combination treatments. A statement on the most promising compounds is thus not possible.

## Conclusions

In summary, most improved IL-2-based compounds are currently developed for the treatment of cancer with some compounds in early phase clinical trials for the treatment of autoimmune diseases. Only limited results of RCTs have been published so far, not allowing a clear conclusion on efficacy of improved IL-2-based compounds. However, the very large number of compounds and registered clinical trials reflects the considerable interest in developing improved IL-2 compounds and commitment to bring these to clinical application.

## Data Availability

The full dataset generated for this study is available upon request from the corresponding authors upon publication of this manuscript in a peer-reviewed journal.

## 7. Author contributions

MR and OB: conception of the study. MR, UK, and DS: data collection. MR, UK, DS, and OB: analysis and interpretation of data. MR and OB: writing of the first draft of the article with critical revision and approval by all authors.

## Acknowledgements

This work was supported by the Swiss National Science Foundation (310030-172978 and 310030-200669; to O.B.), Hochspezialisierte Medizin Schwerpunkt Immunologie (HSM-2-Immunologie; to O.B.), the Clinical Research Priority Program CYTIMM-Z of University of Zurich (to O.B.), a Young Talents in Clinical Research Fellowship and a Project Grant by the Swiss Academy of Medical Sciences and G. & J. Bangerter-Rhyner Foundation (YTCR 32/18 and YTCR 08/20; to M.R.), and a Filling-the-Gap Fellowship by University of Zurich (to M.R.).

## 9. Supplementary material

**Table S1:** Preferred Reporting Items for Systematic Reviews and Meta-Analyses (PRISMA) checklist.

| Section and Topic | Item # | Checklist item | Location where item is reported |
| --- | --- | --- | --- |
| **TITLE** |  |  |  |
| Title | 1 | Identify the report as a systematic review. | Page 1 |
| **ABSTRACT** |  |  |  |
| Abstract | 2 | See the PRISMA 2020 for Abstracts checklist. | Page 2 |
| **INTRODUCTION** |  |  |  |
| Rationale | 3 | Describe the rationale for the review in the context of existing knowledge. | Page 4 |
| Objectives | 4 | Provide an explicit statement of the objective(s) or question(s) the review addresses. | Page 5 |
| **METHODS** |  |  |  |
| Eligibility criteria | 5 | Specify the inclusion and exclusion criteria for the review and how studies were grouped for the syntheses. | Pages 6-7 |
| Information sources | 6 | Specify all databases, registers, websites, organisations, reference lists and other sources searched or consulted to identify studies. Specify the date when each source was last searched or consulted. | Pages 6-7 |
| Search strategy | 7 | Present the full search strategies for all databases, registers and websites, including any filters and limits used. | Pages 6-7 |
| Selection process | 8 | Specify the methods used to decide whether a study met the inclusion criteria of the review, including how many reviewers screened each record and each report retrieved, whether they worked independently, and if applicable, details of automation tools used in the process. | Pages 6-7 |
| Data collection process | 9 | Specify the methods used to collect data from reports, including how many reviewers collected data from each report, whether they worked independently, any processes for obtaining or confirming data from study investigators, and if applicable, details of automation tools used in the process. | Pages 6-7 |
| Data items | 10a | List and define all outcomes for which data were sought. Specify whether all results that were compatible with each outcome domain in each study were sought (e.g. for all measures, time points, analyses), and if not, the methods used to decide which results to collect. | Pages 6-7 |
|  | 10b | List and define all other variables for which data were sought (e.g. participant and intervention characteristics, funding sources). Describe any assumptions made about any missing or unclear information. | Pages 6-7 |
| Study risk of bias assessment | 11 | Specify the methods used to assess risk of bias in the included studies, including details of the tool(s) used, how many reviewers assessed each study and whether they worked independently, and if applicable, details of automation tools used in the process. | Pages 6-7 |
| Effect measures | 12 | Specify for each outcome the effect measure(s) (e.g. risk ratio, mean difference) used in the synthesis or presentation of results. | Pages 6-7 |
| Synthesis methods | 13a | Describe the processes used to decide which studies were eligible for each synthesis (e.g. tabulating the study intervention characteristics and comparing against the planned groups for each synthesis (item #5)). | Pages 6-7 |
|  | 13b | Describe any methods required to prepare the data for presentation or synthesis, such as handling of missing summary statistics, or data conversions. | Pages 6-7 |
|  | 13c | Describe any methods used to tabulate or visually display results of individual studies and syntheses. | Pages 6-7 |
|  | 13d | Describe any methods used to synthesize results and provide a rationale for the choice(s). If meta-analysis was performed, describe the model(s), method(s) to identify the presence and extent of statistical heterogeneity, and software package(s) used. | Pages 6-7 |
|  | 13e | Describe any methods used to explore possible causes of heterogeneity among study results (e.g. subgroup analysis, meta-regression). | Pages 6-7 |
|  | 13f | Describe any sensitivity analyses conducted to assess robustness of the synthesized results. | Pages 6-7 |
| Reporting bias assessment | 14 | Describe any methods used to assess risk of bias due to missing results in a synthesis (arising from reporting biases). | Pages 6-7 |
| Certainty assessment | 15 | Describe any methods used to assess certainty (or confidence) in the body of evidence for an outcome. | Pages 6-7 |
| **RESULTS** |  |  |  |
| Study selection | 16a | Describe the results of the search and selection process, from the number of records identified in the search to the number of studies included in the review, ideally using a flow diagram. | Page 36 |
|  | 16b | Cite studies that might appear to meet the inclusion criteria, but which were excluded, and explain why they were excluded. | Page 8 |
| Study characteristics | 17 | Cite each included study and present its characteristics. | Table 1 and table 2 |
| Risk of bias in studies | 18 | Present assessments of risk of bias for each included study. | Table S2 |
| Results of individual studies | 19 | For all outcomes, present, for each study: (a) summary statistics for each group (where appropriate) and (b) an effect estimate and its precision (e.g. confidence/credible interval), ideally using structured tables or plots. | Table 1 and table 2 |
| Results of syntheses | 20a | For each synthesis, briefly summarise the characteristics and risk of bias among contributing studies. | Pages 8-14 |
|  | 20b | Present results of all statistical syntheses conducted. If meta-analysis was done, present for each the summary estimate and its precision (e.g. confidence/credible interval) and measures of statistical heterogeneity. If comparing groups, describe the direction of the effect. | Table 1 and table 2 |
|  | 20c | Present results of all investigations of possible causes of heterogeneity among study results. | Pages 8-14 |
|  | 20d | Present results of all sensitivity analyses conducted to assess the robustness of the synthesized results. | Pages 8-14 |
| Reporting biases | 21 | Present assessments of risk of bias due to missing results (arising from reporting biases) for each synthesis assessed. | Pages 8-14 |
| Certainty of evidence | 22 | Present assessments of certainty (or confidence) in the body of evidence for each outcome assessed. | Pages 8-14 |
| **DISCUSSION** |  |  |  |
| Discussion | 23a | Provide a general interpretation of the results in the context of other evidence. | Pages 15-16 |
|  | 23b | Discuss any limitations of the evidence included in the review. | Pages 15-16 |
|  | 23c | Discuss any limitations of the review processes used. | Pages 15-16 |
|  | 23d | Discuss implications of the results for practice, policy, and future research. | Pages 15-16 |
| **OTHER INFORMATION** |  |  |  |
| Registration and protocol | 24a | Provide registration information for the review, including register name and registration number, or state that the review was not registered. | Page 6 |
|  | 24b | Indicate where the review protocol can be accessed, or state that a protocol was not prepared. | Materials and methods was prepared as protocol before initiation of review. |
|  | 24c | Describe and explain any amendments to information provided at registration or in the protocol. | NA |
| Support | 25 | Describe sources of financial or non-financial support for the review, and the role of the funders or sponsors in the review. | Page 17 |
| Competing interests | 26 | Declare any competing interests of review authors. | Page 17 |
| Availability of data, code and other materials | 27 | Report which of the following are publicly available and where they can be found: template data collection forms; data extracted from included studies; data used for all analyses; analytic code; any other materials used in the review. | Provided in the review or retrievable from the corresponding author. |
| Reproduced from Ref. (17). |  |  |  |

**Table S2:** Risk of bias assessment for randomized controlled studies reporting clinical results.

|  | Reporting |  |  |  |  |  |  |  |  |  | External validity |  |  | Internal |  |  |  |  |  |  | Source of patients included in the study |  |  |  |  |  | Power |
| --- | --- | --- | --- | --- | --- | --- | --- | --- | --- | --- | --- | --- | --- | --- | --- | --- | --- | --- | --- | --- | --- | --- | --- | --- | --- | --- | --- |
|  | 1 | 2 | 3 | 4 | 5 | 6 | 7 | 8 | 9 | 10 | 11 | 12 | 13 | 14 | 15 | 16 | 17 | 18 | 19 | 20 | 21 | 22 | 23 | 24 | 25 | 26 | 27 |
| NCT01383148 (Quoix et al. 2015) | x | x | x | x | x | x | x | x | x | x | o | o | o | x | x | x | — | x | x | x | x | x | x | x | x | x | x |
| NCT00415818 (Quoix et al. 2011) | x | x | x | x | x | x | x | x | x | x | o | o | o | x | x | x | — | x | x | x | x | x | x | x | x | x | x |
| NCT03635983 (PIVOT IO-001) | o | o | o | o | o | o | o | o | o | o | o | o | o | o | o | o | o | o | o | o | o | o | o | o | o | o | o |
| NCT03729245 (PIVOT-09) | o | o | o | o | o | o | o | o | o | o | o | o | o | o | o | o | o | o | o | o | o | o | o | o | o | o | o |
| NCT03556007 (Fanton et al. 2022) | x | x | x | x | x | x | x | x | x | x | o | o | o | x | x | x | — | x | x | x | x | x | x | x | o | x | x |

## References

1. O. Boyman, J. Sprent, The role of interleukin-2 during homeostasis and activation of the immune system. Nat Rev Immunol 12, 180–190 (2012).

2. A. Yu, I. Snowhite, F. Vendrame, M. Rosenzwajg, D. Klatzmann, A. Pugliese, T. R. Malek, Selective IL-2 responsiveness of regulatory T cells through multiple intrinsic mechanisms supports the use of low-dose IL-2 therapy in type 1 diabetes. Diabetes 64, 2172–2183 (2015).

3. S. A. Rosenberg, IL-2: the first effective immunotherapy for human cancer. J Immunol 192, 5451–5458 (2014).

4. R. Hernandez, J. Poder, K. M. LaPorte, T. R. Malek, Engineering IL-2 for immunotherapy of autoimmunity and cancer. Nat Rev Immunol 22, 614–628 (2022).

5. R. Bright, B. J. Coventry, N. Eardley-Harris, N. Briggs, Clinical Response Rates From Interleukin-2 Therapy for Metastatic Melanoma Over 30 Years’ Experience: A Meta-Analysis of 3312 Patients. J Immunother 40, 21–30 (2017).

6. S. A. Rosenberg, J. C. Yang, D. E. White, S. M. Steinberg, Durability of complete responses in patients with metastatic cancer treated with high-dose interleukin-2: identification of the antigens mediating response. Ann Surg 228, 307–319 (1998).

7. C. Krieg, S. Letourneau, G. Pantaleo, O. Boyman, Improved IL-2 immunotherapy by selective stimulation of IL-2 receptors on lymphocytes and endothelial cells. Proc Natl Acad Sci U S A 107, 11906–11911 (2010).

8. M. E. Raeber, R. A. Rosalia, D. Schmid, U. Karakus, O. Boyman, Interleukin-2 signals converge in a lymphoid-dendritic cell pathway that promotes anticancer immunity. Sci Transl Med 12, eaba5464 (2020).

9. S. Sakaguchi, N. Sakaguchi, M. Asano, M. Itoh, M. Toda, Immunologic self-tolerance maintained by activated T cells expressing IL-2 receptor alpha-chains (CD25). Breakdown of a single mechanism of self-tolerance causes various autoimmune diseases. J Immunol 155, 1151–1164 (1995).

10. J. Y. Humrich, C. von Spee-Mayer, E. Siegert, T. Alexander, F. Hiepe, A. Radbruch, G. R. Burmester, G. Riemekasten, Rapid induction of clinical remission by low-dose interleukin-2 in a patient with refractory SLE. Ann Rheum Dis 74, 791–792 (2015).

11. H. Grasshoff, S. Comduhr, L. R. Monne, A. Muller, P. Lamprecht, G. Riemekasten, J. Y. Humrich, Low-Dose IL-2 Therapy in Autoimmune and Rheumatic Diseases. Front Immunol 12, 648408 (2021).

12. D. Klatzmann, A. K. Abbas, The promise of low-dose interleukin-2 therapy for autoimmune and inflammatory diseases. Nat Rev Immunol 15, 283–294 (2015).

13. J. Koreth, K. Matsuoka, H. T. Kim, S. M. McDonough, B. Bindra, E. P. Alyea, 3rd, P. Armand, C. Cutler, V. T. Ho, N. S. Treister, D. C. Bienfang, S. Prasad, D. Tzachanis, R. M. Joyce, D. E. Avigan, J. H. Antin, J. Ritz, R. J. Soiffer, Interleukin-2 and regulatory T cells in graft-versus-host disease. N Engl J Med 365, 2055–2066 (2011).

14. D. Saadoun, M. Rosenzwajg, F. Joly, A. Six, F. Carrat, V. Thibault, D. Sene, P. Cacoub, D. Klatzmann, Regulatory T-cell responses to low-dose interleukin-2 in HCV-induced vasculitis. N Engl J Med 365, 2067–2077 (2011).

15. J. Y. Humrich, P. Cacoub, M. Rosenzwajg, F. Pitoiset, H. P. Pham, J. Guidoux, D. Leroux, T. Vazquez, G. Riemekasten, J. S. Smolen, G. Tsokos, D. Klatzmann, Low-dose interleukin-2 therapy in active systemic lupus erythematosus (LUPIL-2): a multicentre, double-blind, randomised and placebo-controlled phase II trial. Ann Rheum Dis 81, 1685–1694 (2022).

16. J. He, R. Zhang, M. Shao, X. Zhao, M. Miao, J. Chen, J. Liu, X. Zhang, X. Zhang, Y. Jin, Y. Wang, S. Zhang, L. Zhu, A. Jacob, R. Jia, X. You, X. Li, C. Li, Y. Zhou, Y. Yang, H. Ye, Y. Liu, Y. Su, N. Shen, J. Alexander, J. Guo, J. Ambrus, X. Lin, D. Yu, X. Sun, Z. Li, Efficacy and safety of low-dose IL-2 in the treatment of systemic lupus erythematosus: a randomised, double-blind, placebo-controlled trial. Ann Rheum Dis 79, 141–149 (2020).

17. M. J. Page, J. E. McKenzie, P. M. Bossuyt, I. Boutron, T. C. Hoffmann, C. D. Mulrow, L. Shamseer, J. M. Tetzlaff, E. A. Akl, S. E. Brennan, R. Chou, J. Glanville, J. M. Grimshaw, A. Hrobjartsson, M. M. Lalu, T. Li, E. W. Loder, E. Mayo-Wilson, S. McDonald, L. A. McGuinness, L. A. Stewart, J. Thomas, A. C. Tricco, V. A. Welch, P. Whiting, D. Moher, The PRISMA 2020 statement: an updated guideline for reporting systematic reviews. BMJ 372, n71 (2021).

18. M. E. Raeber, D. Sahin, O. Boyman, Interleukin-2-based therapies in cancer. Sci Transl Med 14, abo5409 (2022).

19. S. H. Downs, N. Black, The feasibility of creating a checklist for the assessment of the methodological quality both of randomised and non-randomised studies of health care interventions. J Epidemiol Community Health 52, 377–384 (1998).

20. U. Sahin, L. Kranz, M. Vormehr, M. Diken, S. Kreiter, B. Tillmann, Treatment using cytokine encoding RNA, patent number EP2019053134W, 02.08.2019.

21. A. M. Nash, M. I. Jarvis, S. Aghlara-Fotovat, S. Mukherjee, A. Hernandez, A. D. Hecht, P. D. Rios, S. Ghani, I. Joshi, D. Isa, Y. Cui, S. Nouraein, J. Z. Lee, C. Xu, D. Y. Zhang, R. A. Sheth, W. Peng, J. Oberholzer, O. A. Igoshin, A. A. Jazaeri, O. Veiseh, Clinically translatable cytokine delivery platform for eradication of intraperitoneal tumors. Sci Adv 8, eabm1032 (2022).

22. E. Quoix, H. Lena, G. Losonczy, F. Forget, C. Chouaid, Z. Papai, R. Gervais, C. Ottensmeier, A. Szczesna, A. Kazarnowicz, J. T. Beck, V. Westeel, E. Felip, D. Debieuvre, A. Madroszyk, J. Adam, G. Lacoste, A. Tavernaro, B. Bastien, C. Halluard, T. Palanche, J. M. Limacher, TG4010 immunotherapy and first-line chemotherapy for advanced non-small-cell lung cancer (TIME): results from the phase 2b part of a randomised, double-blind, placebo-controlled, phase 2b/3 trial. Lancet Oncol 17, 212–223 (2016).

23. E. Quoix, R. Ramlau, V. Westeel, Z. Papai, A. Madroszyk, A. Riviere, P. Koralewski, J. L. Breton, E. Stoelben, D. Braun, D. Debieuvre, H. Lena, M. Buyse, M. P. Chenard, B. Acres, G. Lacoste, B. Bastien, A. Tavernaro, N. Bizouarne, J. Y. Bonnefoy, J. M. Limacher, Therapeutic vaccination with TG4010 and first-line chemotherapy in advanced non-small-cell lung cancer: a controlled phase 2B trial. Lancet Oncol 12, 1125–1133 (2011).

24. R. Dreicer, W. M. Stadler, F. R. Ahmann, T. Whiteside, N. Bizouarne, B. Acres, J. M. Limacher, P. Squiban, A. Pantuck, MVA-MUC1-IL2 vaccine immunotherapy (TG4010) improves PSA doubling time in patients with prostate cancer with biochemical failure. Invest New Drugs 27, 379–386 (2009).

25. C. Rochlitz, R. Figlin, P. Squiban, M. Salzberg, M. Pless, R. Herrmann, E. Tartour, Y. Zhao, N. Bizouarne, M. Baudin, B. Acres, Phase I immunotherapy with a modified vaccinia virus (MVA) expressing human MUC1 as antigen-specific immunotherapy in patients with MUC1-positive advanced cancer. J Gene Med 5, 690–699 (2003).

26. L. Chan, N. Hardwick, D. Darling, J. Galea-Lauri, J. Gaken, S. Devereux, M. Kemeny, G. Mufti, F. Farzaneh, IL-2/B7.1 (CD80) fusagene transduction of AML blasts by a self-inactivating lentiviral vector stimulates T cell responses in vitro: a strategy to generate whole cell vaccines for AML. Mol Ther 11, 120–131 (2005).

27. D. Saltzman, E. Moradian, A Phase 2 Study of Saltikva (Salmonella-IL2) in Metastatic Pancreatic Cancer, poster presented at the 27th Congress of the European Association for Cancer Research, Virtual Meeting, 09.06.2021.

28. K. F. Card, S. A. Price-Schiavi, B. Liu, E. Thomson, E. Nieves, H. Belmont, J. Builes, J. A. Jiao, J. Hernandez, J. Weidanz, L. Sherman, J. L. Francis, A. Amirkhosravi, H. C. Wong, A soluble single-chain T-cell receptor IL-2 fusion protein retains MHC-restricted peptide specificity and IL-2 bioactivity. Cancer Immunol Immunother 53, 345–357 (2004).

29. M. N. Fishman, J. A. Thompson, G. K. Pennock, R. Gonzalez, L. M. Diez, A. I. Daud, J. S. Weber, B. Y. Huang, S. Tang, P. R. Rhode, H. C. Wong, Phase I trial of ALT-801, an interleukin-2/T-cell receptor fusion protein targeting p53 (aa264-272)/HLA-A*0201 complex, in patients with advanced malignancies. Clin Cancer Res 17, 7765–7775 (2011).

30. J. A. Hank, J. E. Surfus, J. Gan, P. Jaeger, S. D. Gillies, R. A. Reisfeld, P. M. Sondel, Activation of human effector cells by a tumor reactive recombinant anti-ganglioside GD2 interleukin-2 fusion protein (ch14.18-IL2). Clin Cancer Res 2, 1951–1959 (1996).

31. S. Shusterman, W. B. London, S. D. Gillies, J. A. Hank, S. D. Voss, R. C. Seeger, C. P. Reynolds, J. Kimball, M. R. Albertini, B. Wagner, J. Gan, J. Eickhoff, K. B. DeSantes, S. L. Cohn, T. Hecht, B. Gadbaw, R. A. Reisfeld, J. M. Maris, P. M. Sondel, Antitumor activity of hu14.18-IL2 in patients with relapsed/refractory neuroblastoma: a Children’s Oncology Group (COG) phase II study. J Clin Oncol 28, 4969–4975 (2010).

32. K. L. Osenga, J. A. Hank, M. R. Albertini, J. Gan, A. G. Sternberg, J. Eickhoff, R. C. Seeger, K. K. Matthay, C. P. Reynolds, C. Twist, M. Krailo, P. C. Adamson, R. A. Reisfeld, S. D. Gillies, P. M. Sondel, G. Children’s Oncology, A phase I clinical trial of the hu14.18-IL2 (EMD 273063) as a treatment for children with refractory or recurrent neuroblastoma and melanoma: a study of the Children’s Oncology Group. Clin Cancer Res 12, 1750–1759 (2006).

33. M. R. Albertini, J. A. Hank, B. Gadbaw, J. Kostlevy, J. Haldeman, H. Schalch, J. Gan, K. Kim, J. Eickhoff, S. D. Gillies, P. M. Sondel, Phase II trial of hu14.18-IL2 for patients with metastatic melanoma. Cancer Immunol Immunother 61, 2261–2271 (2012).

34. S. D. Gillies, Y. Lan, S. Williams, F. Carr, S. Forman, A. Raubitschek, K. M. Lo, An anti-CD20-IL-2 immunocytokine is highly efficacious in a SCID mouse model of established human B lymphoma. Blood 105, 3972–3978 (2005).

35. F. Lansigan, R. Nakamura, D. P. Quick, D. Vlock, A. Raubitschek, S. D. Gillies, V. Bachanova, DI- Leu16-IL2, an Anti-CD20-Interleukin-2 Immunocytokine, Is Safe and Active in Patients with Relapsed and Refractory B-Cell Lymphoma: A Report of Maximum Tolerated Dose, Optimal Biologic Dose, and Recommended Phase 2 Dose. Blood 128, 620–620 (2016).

36. Sanofi, Third quarter 2022 results, https://www.sanofi.com/en/investors/financial-results-and-events/financial-results/Q3-results-2022, accessed on 13.11.2022.

37. B. Carnemolla, L. Borsi, E. Balza, P. Castellani, R. Meazza, A. Berndt, S. Ferrini, H. Kosmehl, D. Neri, L. Zardi, Enhancement of the antitumor properties of interleukin-2 by its targeted delivery to the tumor blood vessel extracellular matrix. Blood 99, 1659–1665 (2002).

38. B. Weide, T. Eigentler, C. Catania, P. A. Ascierto, S. Cascinu, J. C. Becker, A. Hauschild, A. Romanini, R. Danielli, R. Dummer, U. Trefzer, G. Elia, D. Neri, C. Garbe, A phase II study of the L19IL2 immunocytokine in combination with dacarbazine in advanced metastatic melanoma patients. Cancer Immunol Immunother 68, 1547–1559 (2019).

39. M. Johannsen, G. Spitaleri, G. Curigliano, J. Roigas, S. Weikert, C. Kempkensteffen, A. Roemer, C. Kloeters, P. Rogalla, G. Pecher, K. Miller, A. Berndt, H. Kosmehl, E. Trachsel, M. Kaspar, V. Lovato, R. Gonzalez-Iglesias, L. Giovannoni, H. D. Menssen, D. Neri, F. de Braud, The tumour-targeting human L19-IL2 immunocytokine: preclinical safety studies, phase I clinical trial in patients with solid tumours and expansion into patients with advanced renal cell carcinoma. Eur J Cancer 46, 2926–2935 (2010).

40. T. K. Eigentler, B. Weide, F. de Braud, G. Spitaleri, A. Romanini, A. Pflugfelder, R. Gonzalez-Iglesias, A. Tasciotti, L. Giovannoni, K. Schwager, V. Lovato, M. Kaspar, E. Trachsel, H. D. Menssen, D. Neri, C. Garbe, A dose-escalation and signal-generating study of the immunocytokine L19-IL2 in combination with dacarbazine for the therapy of patients with metastatic melanoma. Clin Cancer Res 17, 7732–7742 (2011).

41. E. J. Van Limbergen, A. Hoeben, R. I. Y. Lieverse, R. Houben, C. Overhof, A. Postma, J. Zindler, F. Verhelst, L. J. Dubois, D. De Ruysscher, E. G. C. Troost, P. Lambin, Toxicity of L19-Interleukin 2 Combined with Stereotactic Body Radiation Therapy: A Phase 1 Study. Int J Radiat Oncol Biol Phys 109, 1421–1430 (2021).

42. R. Danielli, R. Patuzzo, A. M. Di Giacomo, G. Gallino, A. Maurichi, A. Di Florio, O. Cutaia, A. Lazzeri, C. Fazio, C. Miracco, L. Giovannoni, G. Elia, D. Neri, M. Maio, M. Santinami, Intralesional administration of L19-IL2/L19-TNF in stage III or stage IVM1a melanoma patients: results of a phase II study. Cancer Immunol Immunother 64, 999–1009 (2015).

43. J. Marlind, M. Kaspar, E. Trachsel, R. Sommavilla, S. Hindle, C. Bacci, L. Giovannoni, D. Neri, Antibody-mediated delivery of interleukin-2 to the stroma of breast cancer strongly enhances the potency of chemotherapy. Clin Cancer Res 14, 6515–6524 (2008).

44. C. Catania, M. Maur, R. Berardi, A. Rocca, A. M. Giacomo, G. Spitaleri, C. Masini, C. Pierantoni, R. Gonzalez-Iglesias, G. Zigon, A. Tasciotti, L. Giovannoni, V. Lovato, G. Elia, H. D. Menssen, D. Neri, S. Cascinu, P. F. Conte, F. Braud, The tumor-targeting immunocytokine F16-IL2 in combination with doxorubicin: dose escalation in patients with advanced solid tumors and expansion into patients with metastatic breast cancer. Cell Adh Migr 9, 14–21 (2015).

45. A. F. Berdel, C. Rollig, M. Wermke, L. Angenendt, L. Ruhnke, J.-H. Mikesch, T. Hemmerle, K. Wethmar, T. Kessler, M. Gerwing, D. Hescheler, M. Schäfers, W. Hartmann, B. Altvater, C. Rossig, G. Lenz, M. Stelljes, B. Rueter, D. Neri, W. E. Berdel, C. Schliemann, A Phase I Trial of the Antibody-Cytokine Fusion Protein F16IL2 in Combination with Anti-CD33 Immunotherapy for Posttransplant AML Relapse. Blood 138, 2345 (2021).

46. C. J. Nirschl, H. R. Brodkin, D. J. Hicklin, N. Ismail, K. Morris, C. Seidel-Dugan, P. Steiner, Z. Steuert, J. M. Sullivan, E. Tyagi, W. M. Winston, A. Salmeron, Discovery of a Conditionally Activated IL-2 that Promotes Antitumor Immunity and Induces Tumor Regression. Cancer Immunol Res 10, 581–596 (2022).

47. A. B. Shanafelt, Y. Lin, M. C. Shanafelt, C. P. Forte, N. Dubois-Stringfellow, C. Carter, J. A. Gibbons, S. L. Cheng, K. A. Delaria, R. Fleischer, J. M. Greve, R. Gundel, K. Harris, R. Kelly, B. Koh, Y. Li, L. Lantz, P. Mak, L. Neyer, M. J. Plym, S. Roczniak, D. Serban, J. Thrift, L. Tsuchiyama, M. Wetzel, M. Wong, A. Zolotorev, A T-cell-selective interleukin 2 mutein exhibits potent antitumor activity and is well tolerated in vivo. Nat Biotechnol 18, 1197–1202 (2000).

48. K. Margolin, M. B. Atkins, J. P. Dutcher, M. S. Ernstoff, J. W. Smith, 2nd, J. I. Clark, J. Baar, J. Sosman, J. Weber, C. Lathia, J. Brunetti, F. Cihon, B. Schwartz, Phase I trial of BAY 50-4798, an interleukin-2-specific agonist in advanced melanoma and renal cancer. Clin Cancer Res 13, 3312–3319 (2007).

49. N. Arenas-Ramirez, J. Woytschak, O. Boyman, Interleukin-2: Biology, Design and Application. Trends Immunol 36, 763–777 (2015).

50. S. D. Gillies, Y. Lan, T. Hettmann, B. Brunkhorst, Y. Sun, S. O. Mueller, K. M. Lo, A low-toxicity IL-2-based immunocytokine retains antitumor activity despite its high degree of IL-2 receptor selectivity. Clin Cancer Res 17, 3673–3685 (2011).

51. S. Gillessen, U. S. Gnad-Vogt, E. Gallerani, J. Beck, C. Sessa, A. Omlin, M. R. Mattiacci, B. Liedert, D. Kramer, J. Laurent, D. E. Speiser, R. Stupp, A phase I dose-escalation study of the immunocytokine EMD 521873 (Selectikine) in patients with advanced solid tumours. Eur J Cancer 49, 35–44 (2013).

52. M. M. van den Heuvel, M. Verheij, R. Boshuizen, J. Belderbos, A. M. Dingemans, D. De Ruysscher, J. Laurent, R. Tighe, J. Haanen, S. Quaratino, NHS-IL2 combined with radiotherapy: preclinical rationale and phase Ib trial results in metastatic non-small cell lung cancer following first-line chemotherapy. J Transl Med 13, 32 (2015).

53. O. Boyman, M. Kovar, M. P. Rubinstein, C. D. Surh, J. Sprent, Selective stimulation of T cell subsets with antibody-cytokine immune complexes. Science 311, 1924–1927 (2006).

54. N. Arenas-Ramirez, C. Zou, S. Popp, D. Zingg, B. Brannetti, E. Wirth, T. Calzascia, J. Kovarik, L. Sommer, G. Zenke, J. Woytschak, C. H. Regnier, A. Katopodis, O. Boyman, Improved cancer immunotherapy by a CD25-mimobody conferring selectivity to human interleukin-2. Sci Transl Med 8, 367ra166 (2016).

55. D. Sahin, N. Arenas-Ramirez, M. Rath, U. Karakus, M. Humbelin, M. van Gogh, L. Borsig, O. Boyman, An IL-2-grafted antibody immunotherapy with potent efficacy against metastatic cancer. Nat Commun 11, 6440 (2020).

56. E. Garralda, G. Alonso, J. Lopez, H. Läubli, E. Calvo, C. Huber, N. Egli, K. Richter, L. Petersen, C. Lanza, S. Jethwa, S. Costanzo, A. Nair, J. Mouton, D. Di Blasi, C. Bucher, ANV419, an IL-2R-beta-gamma targeted antibody-IL-2 fusion protein, induces selective effector cell proliferation in patients with progressed cancer (Poster CT140), poster presented at the 2022 Annual Meeting of the American Association for Cancer Research, New Orleans, LA, 11 April 2022.

57. G.-A. Kim, D.-E. Kim, Y.-J. Kim, K.-M. Han, H.-S. Kim, S.-H. Park, J.-H. Park, E.-J. Roh, G.-T. Wie, J.-Y. Lee, Abstract 689: Strong anti-tumor activity and stability of IL-2/anti IL-2 conjugate SLC- 3010 in preclinical experiments. Cancer Research 81, 689–689 (2021).

58. J. E. Lopes, J. L. Fisher, H. L. Flick, C. Wang, L. Sun, M. S. Ernstoff, J. C. Alvarez, H. C. Losey, ALKS 4230: a novel engineered IL-2 fusion protein with an improved cellular selectivity profile for cancer immunotherapy. J Immunother Cancer 8, e000673 (2020).

59. U. N. Vaishampayan, P. Tomczak, J. Muzaffar, I. S. Winer, S. D. Rosen, C. J. Hoimes, A. Chauhan, A. Spreafico, K. D. Lewis, D. S. Bruno, O. Dumas, D. F. McDermott, J. F. Strauss, Q. S. Chu, L. Gilbert, A. Chaudhry, J. R. Graham, V. Boni, M. S. Ernstoff, V. Velcheti, Nemvaleukin alfa monotherapy and in combination with pembrolizumab in patients (pts) with advanced solid tumors: ARTISTRY-1. Journal of Clinical Oncology 40, 2500–2500 (2022).

60. O. Hamid, S. V. Liu, R. V. Boccia, J. A. Call, T. M. Wise-Draper, A. T. Alistar, J. D. Powderly, B. C. Carthon, U. N. Vaishampayan, A. J. Olszanski, J. M. Wrangle, A. F. Shields, S. A. A. Piha-Paul, M. Monali Desai, Y. Du, P. Lei Sun, Y. Wang, H. Losey, C. Hopkinson, M. S. Ernstoff, Selection of the recommended phase 2 dose (RP2D) for subcutaneous nemvaleukin alfa: ARTISTRY-2. Journal of Clinical Oncology 39, 2552–2552 (2021).

61. D. H. Charych, U. Hoch, J. L. Langowski, S. R. Lee, M. K. Addepalli, P. B. Kirk, D. Sheng, X. Liu, P. W. Sims, L. A. VanderVeen, C. F. Ali, T. K. Chang, M. Konakova, R. L. Pena, R. S. Kanhere, Y. M. Kirksey, C. Ji, Y. Wang, J. Huang, T. D. Sweeney, S. S. Kantak, S. K. Doberstein, NKTR-214, an Engineered Cytokine with Biased IL2 Receptor Binding, Increased Tumor Exposure, and Marked Efficacy in Mouse Tumor Models. Clin Cancer Res 22, 680–690 (2016).

62. S. E. Bentebibel, M. E. Hurwitz, C. Bernatchez, C. Haymaker, C. W. Hudgens, H. M. Kluger, M. T. Tetzlaff, M. A. Tagliaferri, J. Zalevsky, U. Hoch, C. Fanton, S. Aung, P. Hwu, B. D. Curti, N. M. Tannir, M. Sznol, A. Diab, A First-in-Human Study and Biomarker Analysis of NKTR-214, a Novel IL2Rbetagamma-Biased Cytokine, in Patients with Advanced or Metastatic Solid Tumors. Cancer Discov 9, 711–721 (2019).

63. Nektar Therapeutics and Bristol Myers Squibb, Bristol Myers Squibb and Nektar Announce Update on Phase 3 PIVOT IO-001 Trial Evaluating Bempegaldesleukin (BEMPEG) in Combination with Opdivo (nivolumab) in Previously Untreated Unresectable or Metastatic Melanoma, press release published on Business Wire, 14.03.2022.

64. Nektar Therapeutics, Bristol Myers Squibb, Nektar and Bristol Myers Squibb Announce Update on Clinical Development Program for Bempegaldesleukin (BEMPEG) in Combination with Opdivo (nivolumab), press release published on Cision PR Newswire, 14.04.2022.

65. A. Diab, S. S. Tykodi, G. A. Daniels, M. Maio, B. D. Curti, K. D. Lewis, S. Jang, E. Kalinka, I. Puzanov, A. I. Spira, D. C. Cho, S. Guan, E. Puente, T. Nguyen, U. Hoch, S. L. Currie, W. Lin, M. A. Tagliaferri, J. Zalevsky, M. Sznol, M. E. Hurwitz, Bempegaldesleukin Plus Nivolumab in First-Line Metastatic Melanoma. J Clin Oncol 39, 2914–2925 (2021).

66. A. Diab, B. Curti, M. Bilen, A. Brohl, E. Domingo-Musibay, E. Borazanci, C. Fanton, C. Haglund, M. Vimal, M. Muhsin, M. Marcondes, A. Nguyen, M. Tagliaferri, W. Lin, J. Zalevsky, S. D’Angelo, 368 REVEAL: Phase 1 dose-escalation study of NKTR-262, a novel TLR7/8 agonist, plus bempegaldesleukin: local innate immune activation and systemic adaptive immune expansion for treating solid tumors. Journal for ImmunoTherapy of Cancer 8, A224–A225 (2020).

67. D. B. Rosen, A. M. Kvarnhammar, B. Laufer, T. Knappe, J. J. Karlsson, E. Hong, Y. C. Lee, D. Thakar, L. A. Zuniga, K. Bang, S. S. Sabharwal, K. Uppal, J. D. Olling, K. Kjaergaard, T. Kurpiers, M. Schnabel, D. Reich, P. Glock, J. Zettler, M. Krusch, A. Bernhard, S. Heinig, V. Konjik, T. Wegge, Y. Hehn, S. Killian, L. Viet, J. Runz, F. Faltinger, M. Tabrizi, K. L. Abel, V. M. Breinholt, S. M. Singel, K. Sprogoe, J. Punnonen, TransCon IL-2 beta/gamma: a novel long-acting prodrug with sustained release of an IL-2Rbeta/gamma-selective IL-2 variant with improved pharmacokinetics and potent activation of cytotoxic immune cells for the treatment of cancer. J Immunother Cancer 10, e004991 (2022).

68. J. L. Ptacin, C. E. Caffaro, L. Ma, K. M. San Jose Gall, H. R. Aerni, N. V. Acuff, R. W. Herman, Y. Pavlova, M. J. Pena, D. B. Chen, L. K. Koriazova, L. K. Shawver, I. B. Joseph, M. E. Milla, An engineered IL-2 reprogrammed for anti-tumor therapy using a semi-synthetic organism. Nat Commun 12, 4785 (2021).

69. F. Janku, R. Abdul-Karim, A. Azad, J. Bendell, G. Falchook, H. K. Gan, T. Tan, J. S. Wang, C. E. Chee, L. Ma, J. Mooney, N. Marina, G. Abbadessa, M. Milla, T. Meniawy, Abstract LB041: THOR- 707 (SAR444245), a novel not-alpha IL-2 as monotherapy and in combination with pembrolizumab in advanced/metastatic solid tumors: Interim results from HAMMER, an open-label, multicenter phase 1/2 Study. Cancer Research 81, LB041–LB041 (2021).

70. U. Sahin, M. Vormehr, L. Kranz, S. Fellermeier-Kopf, A. Muik, F. Gieseke, B. Tillmann, S. Witzel, IL-2 Agonists, patent number WO2020020783A1, 30.01.2020.

71. R. Merchant, C. Galligan, M. A. Munegowda, L. B. Pearce, P. Lloyd, P. Smith, F. Merchant, M. D. To, Fine-tuned long-acting interleukin-2 superkine potentiates durable immune responses in mice and non-human primate. J Immunother Cancer 10, e003155 (2022).

72. D. A. Silva, S. Yu, U. Y. Ulge, J. B. Spangler, K. M. Jude, C. Labao-Almeida, L. R. Ali, A. Quijano-Rubio, M. Ruterbusch, I. Leung, T. Biary, S. J. Crowley, E. Marcos, C. D. Walkey, B. D. Weitzner, F. Pardo-Avila, J. Castellanos, L. Carter, L. Stewart, S. R. Riddell, M. Pepper, G. J. L. Bernardes, M. Dougan, K. C. Garcia, D. Baker, De novo design of potent and selective mimics of IL-2 and IL-15. Nature 565, 186–191 (2019).

73. L. Codarri Deak, V. Nicolini, M. Hashimoto, M. Karagianni, P. C. Schwalie, L. Lauener, E. M. Varypataki, M. Richard, E. Bommer, J. Sam, S. Joller, M. Perro, F. Cremasco, L. Kunz, E. Yanguez, T. Husser, R. Schlenker, M. Mariani, V. Tosevski, S. Herter, M. Bacac, I. Waldhauer, S. Colombetti, X. Gueripel, S. Wullschleger, M. Tichet, D. Hanahan, H. T. Kissick, S. Leclair, A. Freimoser-Grundschober, S. Seeber, V. Teichgraber, R. Ahmed, C. Klein, P. Umana, PD-1-cis IL-2R agonism yields better effectors from stem-like CD8(+) T cells. Nature 610, 161–172 (2022).

74. S. N. Quayle, N. Girgis, D. R. Thapa, Z. Merazga, M. M. Kemp, A. Histed, F. Zhao, M. Moreta, P. Ruthardt, S. Hulot, A. Nelson, L. D. Kraemer, D. R. Beal, L. Witt, J. Ryabin, J. Soriano, M. Haydock, E. Spaulding, J. F. Ross, P. A. Kiener, S. Almo, R. Chaparro, R. Seidel, A. Suri, S. Cemerski, K. J. Pienta, M. E. Simcox, CUE-101, a Novel E7-pHLA-IL2-Fc Fusion Protein, Enhances Tumor Antigen-Specific T-Cell Activation for the Treatment of HPV16-Driven Malignancies. Clin Cancer Res 26, 1953–1964 (2020).

75. C. H. Chung, A. D. Colevas, D. Adkins, M. K. Gibson, C. P. Rodriguez, A. Sukari, J. E. Bauman, L. J. Wirth, F. M. Johnson, N. F. Saba, B. Burtness, L. Dunn, T. Y. Seiwert, F. P. Worden, J. Muzaffar, S. Margossian, R. Moniz, S. N. Quayle, M. Levisetti, S. I. Pai, A phase 1 dose-escalation and expansion study of CUE-101, a novel HPV16 E7-pHLA-IL2-Fc fusion protein, given alone and in combination with pembrolizumab in patients with recurrent/metastatic HPV16+ head and neck cancer. Journal of Clinical Oncology 40, 6045–6045 (2022).

76. K.-H. Pyo, Y. J. Koh, C.-B. Synn, J. H. Kim, Y. Byeon, H. N. Jo, Y. S. Kim, W. Lee, D. H. Kim, S. Lee, D. K. Kim, E. j. Lee, B.-C. Ahn, M. H. Hong, M. H. Jang, S. M. Lim, H. R. Kim, S. Y. Nam, B.C. Cho, Abstract 1826: Comprehensive preclinical study on GI-101, a novel CD80-IgG4-IL2 variant protein, as a therapeutic antibody candidate with bispecific immuno-oncology target, poster presented at the 2021 Annual Meeting of the American Association of Cancer Research, Virtual Meeting, 10 April 2021.

77. I. Waldhauer, V. Gonzalez-Nicolini, A. Freimoser-Grundschober, T. K. Nayak, L. Fahrni, R. J. Hosse, D. Gerrits, E. J. W. Geven, J. Sam, S. Lang, E. Bommer, V. Steinhart, E. Husar, S. Colombetti, E. Van Puijenbroek, M. Neubauer, J. M. Cline, P. K. Garg, G. Dugan, F. Cavallo, G. Acuna, J. Charo, V. Teichgraber, S. Evers, O. C. Boerman, M. Bacac, E. Moessner, P. Umana, C. Klein, Simlukafusp alfa (FAP-IL2v) immunocytokine is a versatile combination partner for cancer immunotherapy. MAbs 13, 1913791 (2021).

78. A. Italiano, L. Verlingue, H. Prenen, E. M. Guerra, D. Tosi, R. Perets, I. Lugowska, V. Moiseenko, M. Gumus, C. Arslan, C. R. Lindsay, M. Richard, D. Dejardin, C. Boetsch, A. Kraxner, S. Evers, T. Vardar, N. Keshelava, V. Teichgräber, R. Dziadziuszko, Clinical activity and safety of simlukafusp alfa, an engineered interleukin-2 variant targeted to fibroblast activation protein-α, combined with atezolizumab in patients with recurrent or metastatic cervical cancer. Journal of Clinical Oncology 39, 5510–5510 (2021).

79. A. R. Hansen, C. A. Gomez-Roca, M. P. Lolkema, L. Verlingue, A. Italiano, J. Spicer, N. Steeghs, J. E. Bauman, J. Fayette, J. Niu, H. Prenen, D. Dejardin, C. Boetsch, A. Kraxner, S. Evers, T. Vardar, N. Keshelava, V. Teichgräber, M. Bonomi, 906P Simlukafusp α and cetuximab combination in patients with recurrent, unresectable or metastatic squamous cell carcinoma of the head and neck. Annals of Oncology 32, S805 (2021).

80. J. L. Perez-Gracia, A. R. Hansen, R. H. L. Eefsen, C. A. Gomez-Roca, S. Negrier, P. Pedrazzoli, J.-L. Lee, T. A. Gordoa, C. S. Rodriguez, B. Mellado, V. Moreno, A. Rodriguez-Vida, A. Hussain, N. Getzmann, D. Dejardin, C. Boetsch, A. Kraxner, T. Vardar, V. Teichgräber, T. Powles, Randomized phase Ib study to evaluate safety, pharmacokinetics and therapeutic activity of simlukafusp α in combination with atezolizumab ± bevacizumab in patients with unresectable advanced/ metastatic renal cell carcinoma (RCC) (NCT03063762). Journal of Clinical Oncology 39, 4556–4556 (2021).

81. J. O’Neil, W. Guzman, O. Yerov, P. Johnson, M. Fanny, J. Greene, M. McLaughlin, K. Jenkins, R. O’Donnell, H. Qiu, B. Nicholson, W. Avery, R. C. O’Hagan, Tumor-selective activity of XTX202, a protein-engineered IL-2, in mice without peripheral toxicities in nonhuman primates, poster presented at the 2021 Annual Meeting of the American Society of Clinical Oncology, Virtual Meeting, 04 June 2021.

82. N. Dixit, C. Fanton, J. L. Langowski, Y. Kirksey, P. Kirk, T. Chang, J. Cetz, V. Dixit, G. Kim, P. Kuo, M. Maiti, Y. Tang, L. A. VanderVeen, P. Zhang, M. Lee, J. Ritz, Y. Kamihara, C. Ji, W. Rubas, T. D. Sweeney, S. K. Doberstein, J. Zalevsky, NKTR-358: A novel regulatory T-cell stimulator that selectively stimulates expansion and suppressive function of regulatory T cells for the treatment of autoimmune and inflammatory diseases. J Transl Autoimmun 4, 100103 (2021).

83. OncBioMune, OncBioMune Meets Primary Objective in Trial of ProscaVax Immunotherapy Vaccine for Prostate Cancer, press release published on OncBioMune, 14.09.2017.

84. S. Mitra, A. M. Ring, S. Amarnath, J. B. Spangler, P. Li, W. Ju, S. Fischer, J. Oh, R. Spolski, K. Weiskopf, H. Kohrt, J. E. Foley, S. Rajagopalan, E. O. Long, D. H. Fowler, T. A. Waldmann, K. C. Garcia, W. J. Leonard, Interleukin-2 activity can be fine tuned with engineered receptor signaling clamps. Immunity 42, 826–838 (2015).

85. E. Felip, D. Spigel, O. J. Juan-Vidal, J. T. Beck, C. Aguado de la Rosa, D. Johnson, E. Spath-Schwalbe, A. Reinacher-Schick, D. Rodriguez Abreu, M. Altan, N. Reinmuth, M. Edelman, M. Reck, Q. Zhao, S. Banerjee, S. Chaudhry, D. Chien, M. Tagliaferri, J. Zalevsky, A. Ganti, Preliminary Results from PROPEL: A Phase 1/2 Study of Bempegaldesleukin (BEMPEG: NKTR-214) Plus Pembrolizumab With or Without Chemotherapy in Patients with Metastatic NSCLC, poster presented at the ESMO Immuno-Oncology Congress 2021, Virtual Meeting, 8–11 December 2021.

86. A. M. Levin, D. L. Bates, A. M. Ring, C. Krieg, J. T. Lin, L. Su, I. Moraga, M. E. Raeber, G. R. Bowman, P. Novick, V. S. Pande, C. G. Fathman, O. Boyman, K. C. Garcia, Exploiting a natural conformational switch to engineer an interleukin-2 ’superkine’. Nature 484, 529–533 (2012).

87. C. Klein, I. Waldhauer, V. G. Nicolini, A. Freimoser-Grundschober, T. Nayak, D. J. Vugts, C. Dunn, M. Bolijn, J. Benz, M. Stihle, S. Lang, M. Roemmele, T. Hofer, E. van Puijenbroek, D. Wittig, S. Moser, O. Ast, P. Brunker, I. H. Gorr, S. Neumann, M. C. de Vera Mudry, H. Hinton, F. Crameri, J. Saro, S. Evers, C. Gerdes, M. Bacac, G. van Dongen, E. Moessner, P. Umana, Cergutuzumab amunaleukin (CEA-IL2v), a CEA-targeted IL-2 variant-based immunocytokine for combination cancer immunotherapy: Overcoming limitations of aldesleukin and conventional IL-2-based immunocytokines. Oncoimmunology 6, e1277306 (2017).

88. E. van Brummelen, U. Lassen, I. Melero, J. Tabernero, K. Homicsko, E. Angevin, V. Teichgräber, L. Jukofsky, E. Rossmann, G. Babitzki, A. Patricia Silva, M. Canamero, C. Boetsch, S. Evers, J. Charo, G. Argiles, 4747 - Pharmacokinetics (PK) and Pharmacodynamics (PD) of cergutuzumab amunaleukin (CA), a carcinoembryonic antigen (CEA)-targeted interleukin 2 variant (IL2v) with abolished binding to CD25. Annals of Oncology 28, v403–v427 (2017).

89. C. Fanton, R. Furie, V. Chindalore, R. Levin, I. Diab, N. Dixit, C. Haglund, J. Gibbons, N. Hanan, D. Dickerson, J. Zalevsky, B. L. Kotzin, Selective expansion of regulatory T cells by NKTR-358 in healthy volunteers and patients with systemic lupus erythematosus. J Transl Autoimmun 5, 100152 (2022).

90. L. B. Peterson, C. J. M. Bell, S. K. Howlett, M. L. Pekalski, K. Brady, H. Hinton, D. Sauter, J. A. Todd, P. Umana, O. Ast, I. Waldhauer, A. Freimoser-Grundschober, E. Moessner, C. Klein, R. J. Hosse, L. S. Wicker, A long-lived IL-2 mutein that selectively activates and expands regulatory T cells as a therapy for autoimmune disease. J Autoimmun 95, 1–14 (2018).

91. R. Varma, K. Liu, K. Avery, R. Rashid, S. Schubbert, I. Leung, C. Bonzon, L. Bogaert, J. Desjarlais, M. J. Bernett, Regulatory T Cell Selective IL-2-Fc Fusion Proteins for the Treatment of Autoimmune Diseases. Blood 132, 3709–3709 (2018).

92. J. Visweswaraiah, E. R. Sampson, T. Kiprono, P. Petaipimol, A. Monsef, K. Kis-Toth, K. L. Otipoby, J. Sundy, N. Higginson-Scott, J. L. Viney, Generation of PT101, a highly selective IL-2 mutein for treatment of autoimmune diseases. The Journal of Immunology 206, 66.14–66.14 (2021).

93. A. Ghelani, D. Bates, K. Conner, M. Z. Wu, J. Lu, Y. L. Hu, C. M. Li, A. Chaudhry, S. J. Sohn, Defining the Threshold IL-2 Signal Required for Induction of Selective Treg Cell Responses Using Engineered IL-2 Muteins. Front Immunol 11, 1106 (2020).

94. N. Tchao, K. S. Gorski, T. Yuraszeck, S. J. Sohn, K. Ishida, H. Wong, K. Park, Amg 592 Is an Investigational IL-2 Mutein That Induces Highly Selective Expansion of Regulatory T Cells. Blood 130, 696 (2017).

95. N. Tchao, H. Amouzadeh, N. Sarkar, V. Chow, X. G. Hu, M. Kroenke, H. Wang, R. Zhang, K. Gorski, R. Furie, A. Kivitz, S. Cohen, Efavaleukin Alfa, a Novel IL-2 Mutein, Selectively Expands Regulatory T Cells in Patients with SLE: Interim Results of a Phase 1b Multiple Ascending Dose Study. Arthritis & Rheumatology 73, 3613–3615 (2021).

96. Y. T. Hsieh, C. Hubeau, V. Massa, W. Li, S. Frei, B. Capraro, A. Umana, A. Aherrera, Y. Li, J. Xu, L. Rui, Emerging best-in-class IL-2 variant highlights Treg-directed therapy for autoimmune disease. Annals of the Rheumatic Diseases 79, 195–195 (2020).

